# Whole-genome sequencing characterizes monogenic and polygenic contributions to structural kidney and urinary tract malformations

**DOI:** 10.1101/2024.10.10.24315242

**Authors:** Melanie MY Chan, Omid Sadeghi-Alavijeh, Seth du Preez, Catalin D Voinescu, Loes FM van der Zanden, Sander Groen in ’t Woud, Michiel F Schreuder, Wout Feitz, Enrico Mingardo, Alina C Hilger, Heiko Reutter, Lisanne M Vendrig, Rik Westland, Glenda M Beaman, William G Newman, Adrian S Woolf, Horia C Stanescu, Adam P Levine, Detlef Böckenhauer, Daniel P Gale

## Abstract

**Introduction:** Congenital anomalies of the kidneys and urinary tract (CAKUT) are the commonest cause of kidney failure in children and young adults. Over 50 monogenic causes have been identified, however fewer than 20% of patients have a genetic diagnosis identified using targeted or whole exome sequencing. We sought to characterise the genomic architecture of CAKUT using whole genome sequencing (WGS).

**Methods:** Using WGS data from 1,052 unrelated individuals with CAKUT recruited to the UK’s 100,000 Genomes Project, we used a panel-based framework to determine diagnostic yield and looked for gene-based enrichment of rare variants exome-wide. We performed sequencing based genome-wide association studies (seqGWAS) and estimated the heritability attributable to common and low-frequency variants.

**Results:** The diagnostic yield was 4.9%, increasing to 7.4% in those with kidney agenesis/hypodysplasia and 11.1% in cystic kidney dysplasia. Family history (P=0.02; OR 2.2; 95% CI 1.1-4.4), consanguinity (P=0.01; OR 3.0; 95%CI 1.2-6.9) and extra-renal features (P=1.1×10^-4^; OR 3.1; 95% CI 1.7-5.7) independently predicted a monogenic diagnosis. Exome-wide rare variant and genome-wide common and low-frequency variant (minor allele frequency [MAF] ≥ 0.5%) association testing in a subset of 813 patients and 25,205 ancestry-matched controls identified significant association at 6q16.3 (rs117473527; P=4.83×10^-8^; OR 3.13; 95% CI 2.08-4.72; MAF 0.01) requiring replication. Common and low-frequency variants were estimated to explain 23% (95% CI 1-45%) of the phenotypic variance observed in CAKUT although this was associated with wide confidence intervals. A genomic risk score for posterior urethral valves was also validated in an independent European cohort of 77 cases and 2,746 controls (P < 0.001).

**Conclusions:** Only a minority of patients in this cohort received a monogenic diagnosis, with common and low-frequency variants hypothesised to account for some of this missing heritability. This suggests non-Mendelian genomic factors may be important for the pathogenesis of CAKUT.

**Lay Summary:** This study shows that single-gene causes of isolated and non-familial CAKUT are infrequent, and that genomic testing should be targeted towards those with kidney cysts and/or small kidneys that have not formed properly in the womb. Individuals with a close relative with CAKUT and those with involvement of other organ systems were more likely to receive a genetic diagnosis. These data support a possible polygenic basis for CAKUT, where many common DNA changes cumulatively affect risk, particularly in posterior urethral valves, the most common cause of kidney failure in boys. Larger collaborative genomic studies are needed to increase our ability to identify these DNA changes and the mechanisms and pathways important for kidney and urinary tract development.

## Introduction

Congenital Anomalies of the Kidneys and Urinary Tract (CAKUT) is a widely used diagnostic category encompassing a diverse range of structural kidney and urinary tract malformations. CAKUT are the commonest cause of chronic kidney disease (CKD) in children, accounting for ∼50% of young people requiring dialysis or a kidney transplant.^1,2^ This heterogenous group of malformations are caused by defects in embryonic development and include parenchymal defects (e.g., kidney agenesis, kidney hypodysplasia, cystic dysplasia), collecting system defects (e.g., pelvi-ureteral junction obstruction, primary megaureter, vesicoureteral reflux [VUR]), abnormalities of the lower urinary tract causing bladder dysfunction and/or obstruction (e.g., posterior urethral valves [PUV]) and bladder-exstrophy-epispadias-complex.

CAKUT displays familial clustering^3–5^ with CAKUT-associated syndromes and single gene mouse models implicating a monogenic basis. Over 50 monogenic causes have been reported^6^ with an enrichment of rare copy number variation (CNV) suggesting gene dosage is also important.^7–9^ Many of these genes encode transcription factors, implicating disruption of transcriptional networks as a fundamental mechanism underlying these conditions.

However, several observations suggest CAKUT does not often result from rare single gene defects. First, fewer than 20% of affected individuals have a monogenic cause identified,^10–18^ with diagnostic yield strongly influenced by the cohort studied and testing approach employed. Second, CAKUT demonstrate significant genotype-phenotype heterogeneity, incomplete penetrance and variable expressivity meaning the same variant in a gene can be present in individuals with very different malformations. Third, common variation has previously been associated with susceptibility to PUV,^19^ VUR^20^ and bladder exstrophy.^21^ Finally, maternal diabetes^22^ and obesity^23–25^ have been linked with an increased risk of CAKUT highlighting the potential contribution of genotype-environment interactions to disease risk.^26^ Taken together, these observations suggest a more complex genomic basis underlying the pathogenesis of CAKUT.

Whole-genome sequencing (WGS) enables the comprehensive assessment of variation across the allele frequency spectrum, providing uniform genome-wide coverage of coding and non-coding regions as well as improved detection of disease-causing and structural variants. ^27,28^ Here, we use WGS data to determine diagnostic utility using a virtual panel in 1,052 individuals with CAKUT recruited to the 100,000 Genomes Project.^29^ In addition, our unbiased genome-wide approach interrogating both rare and common variant associations across the entire cohort, and stratified by phenotype, provides support for a polygenic contribution to these phenotypically diverse structural malformations.

## Methods

### The 100,000 Genomes Project

The 100,000 Genomes Project (100KGP) dataset (version 19) consists of WGS data, clinical phenotypes encoded using Human Phenotype Ontology (HPO) codes, and retrospective and prospectively ascertained hospital records for 90,173 individuals with cancer or rare disease, as well as their unaffected relatives.^29^ Full details on the cohort, WGS, alignment, variant calling, and quality control filtering are provided in the Supplementary Methods. The study workflow is shown in Supplementary Figure S1.

### Cohort selection

Patients were recruited to the 100KGP as part of the ‘Congenital anomalies of the kidneys and urinary tract (CAKUT)’ cohort with the following inclusion criteria: CAKUT with syndromic manifestations in other organ systems; isolated CAKUT with a first-degree relative with CAKUT or unexplained CKD; multiple distinct kidney/urinary tract anomalies; CAKUT with unexplained end-stage kidney disease before the age of 25 years. Those with a known genetic or chromosomal abnormality were excluded. Pre-screening for HNF1B and SALL1 was recommended with a personal or family history of diabetes, or imperforate anus, ear or thumb abnormalities, respectively, and only recruited if testing was negative.

For the association analyses, a subset of 813 unrelated patients with GRCh38 aligned genomes were stratified by HPO term (Supplementary Table S1) into the following six groups: kidney anomalies (n=237), cystic renal dysplasia (n=112), obstructive uropathy (excluding PUV; n=177), VUR (n=174), PUV (n=132) and bladder exstrophy (n=97). Unaffected relatives of non-kidney disease participants from the 100KGP, excluding those with SNOMED-CT or ICD-10 codes consistent with kidney disease/failure (Supplementary Table S2) were used as controls. A cohort of unrel ated ancestry-matched cases and controls was generated to minimize the effects of population stratification in this mixed-ancestry cohort (Supplementary Methods and Supplementary Figure S2).

### Identification of disease-causing variants

Probands were assessed using the Genomics England clinical interpretation pipeline to determine a genomic diagnosis with results reviewed by a multi-disciplinary team to determine pathogenicity according to American College of Medical Genetics and Genomics (ACMG)^30^ criteria (see Supplementary Methods). In those without a molecular diagnosis, the top five ranked Exomiser^31^ variants and CNV calls across 286 expertly curated kidney disease genes and loci (CAKUT panel v1.178 and unexplained kidney failure panel v12.7) were manually reviewed.

### Rare variant gene-based analysis

Rare coding single-nucleotide variants (SNVs) and indels with MAF < 0.01% in gnomAD^32^ were aggregated by gene and association testing carried out using SKAT-O^33^ implemented by SAIGE-GENE+^34^ (version 1.3.6). Loss-of-function (LoF) and LoF+missense masks were used. ‘High confidence’ loss-of-function variants (stop gain, splice site and frameshift) were determined by LOFTEE^32^ and missense variants filtered using REVEL^35^ score ≥ 0.75. Variant and sample quality control is detailed in the Supplementary Methods. Sex and the top ten principal components were used as covariates. A Bonferroni adjusted P value of 0.05/number of genes tested was used to determine exome-wide significance.

### seqGWAS

A WGS genome-wide association study (seqGWAS) of SNVs/indels with MAF ≥ 0.5% was performed with SAIGE^36^ (version 1.0.7). seqGWAS was first applied to 813 CAKUT cases and 25,205 ancestry-matched controls and then stratified by phenotype group and ancestry (Supplementary Methods). Sex and the top ten principal components were used as covariates. The conventional genome-wide significance threshold of 5×10^-8^ was employed.

### Meta-Analysis

GWAS summary statistics from the 100KGP obstructive uropathy cohort (177 cases and 24,451 controls) were meta-analysed with the congenital obstructive uropathy cohort in FinnGen^37^ (Release 10; 550 cases and 410,449 controls) using the inverse-variance weighted approach implemented by METAL.^38^ 11,647,818 variants with minor allele count (MAC) > 5 were included. Lambda was 1.01 indicating good control of population stratification. The same approach was used to meta-analyse 4,696,412 variants with MAF > 1% in the 100KGP bladder exstrophy cohort (97 cases and 22,037 controls) with 628 European cases and 7,352 controls from Mingardo et al.^21^ with lambda of 0.98.

### Heritability

Narrow-sense heritability (h^2^) was estimated using three different approaches (GREML-LDMS^39^, LDAK-SumHer^40^, LD-Score Regression (v1.0.1)^41^) in a subset with genetically defined European ancestry (623 cases and 20,060 controls). Observed heritability was transformed to a liability threshold model^42^ adjusting for case:control ratio and an estimated population prevalence of 0.2%.^43^ LDSC^44^ was used to estimate enrichment of functional annotations using the European CAKUT seqGWAS summary statistics and 1000 Genomes (Phase 3) reference panel for LD.

### PUV Genomic Risk Score

PRS-CS (version 1.1.0)^45^ was used to generate a genomic risk score from European male PUV GWAS summary statistics (89 cases and 8,303 controls) using the UK BioBank European LD reference panel. This was applied with PGS Catalog Calculator (version 2.2.0)^46^ to an independent cohort of 77 cases from the Dutch AGORA biobank and 2,746 controls from Genomics England using ten principal components as covariates, normalising to Human Genome Diversity Panel and 1000 Genomes Project scores.

## Results

A total of 1,052 probands with CAKUT were recruited to the 100KGP: 30.2% as singletons, 29.8% duos and 31.0% trios. Table 1 summarizes their clinical and demographic data. The cohort had a median age of 20 years. 13.6% had an affected first-degree relative, 6.1% had reported consanguinity and 28.5% had extra-renal manifestations. The most frequently reported HPO terms in the cohort were hydronephrosis (27.2%), vesicoureteral reflux (VUR) (23.7%) and abnormality of the bladder (20.1%) (Supplementary Figure S3).

**Table 1.** Demographics and clinical characteristics of the CAKUT cohort (n=1,052). Some patients had more than one phenotype. VUR, vesico-ureteral reflux; PUV, posterior urethral valves.

|  |  |
| --- | --- |
| **Median age in years (range)** | 20 (7-86) |
| **Males (%)** | 662 (62.9) |
| **Self-reported ethnicity** |  |
| White (%) | 653 (62.1) |
| South Asian (%) | 98 (9.3) |
| Mixed/Other (%) | 59 (5.6) |
| Black (%) | 28 (2.7) |
| Unknown (%) | 213 (20.2) |
| **Phenotype** |  |
| Kidney anomaly (%) | 393 (37.6) |
| Obstructive uropathy (%) | 341 (32.4) |
| VUR (%) | 249 (23.7) |
| PUV (%) | 173 (16.4) |
| Cystic dysplasia (%) | 162 (15.4) |
| Bladder exstrophy (%) | 108 (10.3) |
| Duplex kidney (%) | 57 (5.4) |
| Ectopic kidney (%) | 39 (3.7) |
| **Extrarenal manifestations (%)** | 300 (28.5) |
| Neurodevelopmental delay (%) | 72 (6.8) |
| Skeletal anomalies (%) | 40 (3.8) |
| Hearing impairment (%) | 31 (2.9) |
| Endocrine (%) | 22 (2.1) |
| Eye anomalies (%) | 22 (2.1) |
| Respiratory (%) | 22 (2.1) |
| Cardiac anomalies (%) | 22 (2.1) |
| **Family history of CAKUT (%)** | **143 (13.6)** |
| **Reported consanguinity (%)** | **64 (6.1)** |
| **Kidney failure (%)** | **285 (27.1)** |
| **Median age of kidney failure in years (range)** | **12 (0-70)** |

### Monogenic structural kidney and urinary tract malformations account for a minority of cases

We first assessed the diagnostic utility of a virtual panel applied to WGS data in this selective CAKUT cohort recruited to the 100KGP. Overall, 4.9% (52/1052) had a genomic diagnosis (Supplementary Table S3), with 71.2% (37/52) of the diagnoses fully explaining the CAKUT phenotype. Excluding individuals with isolated PUV and bladder exstrophy (where monogenic causes are not well established), the diagnostic yield was 6.7% (52/771).

40.4% (21/52) of those with a genomic diagnosis had a pathogenic (P) or likely pathogenic (LP) variant in a known CAKUT gene, most frequently in HNF1B (seven individuals including four with 17q12 deletions; 0.7% of the total cohort). Additional disease-causing variants were reported in PAX2 (3), PBX1 (2), TBX6 (two individuals including one with 16p11.2 microdeletion), WT1 (2) and GREB1L (1) (Figure 1a; Supplementary Table S3). 26.9% (14/52) of the identified P/LP variants affected kidney disease genes which are not usually associated with a CAKUT phenotype: PKD1 (5), PKHD1 (2), CEP290 (2), NPHP1 (2), NPHP3 (1), COL4A5 (1), CUBN (1), SLC3A1 (1) and CLCN5 (1). These likely represent phenocopies in the case of cystic disease, or possible misclassification during recruitment. An additional 23% (12/52) had a monogenic cause for a non CAKUT-associated syndrome identified which did not fully explain the observed kidney and/or urinary tract anomalies.

**Figure 1.**
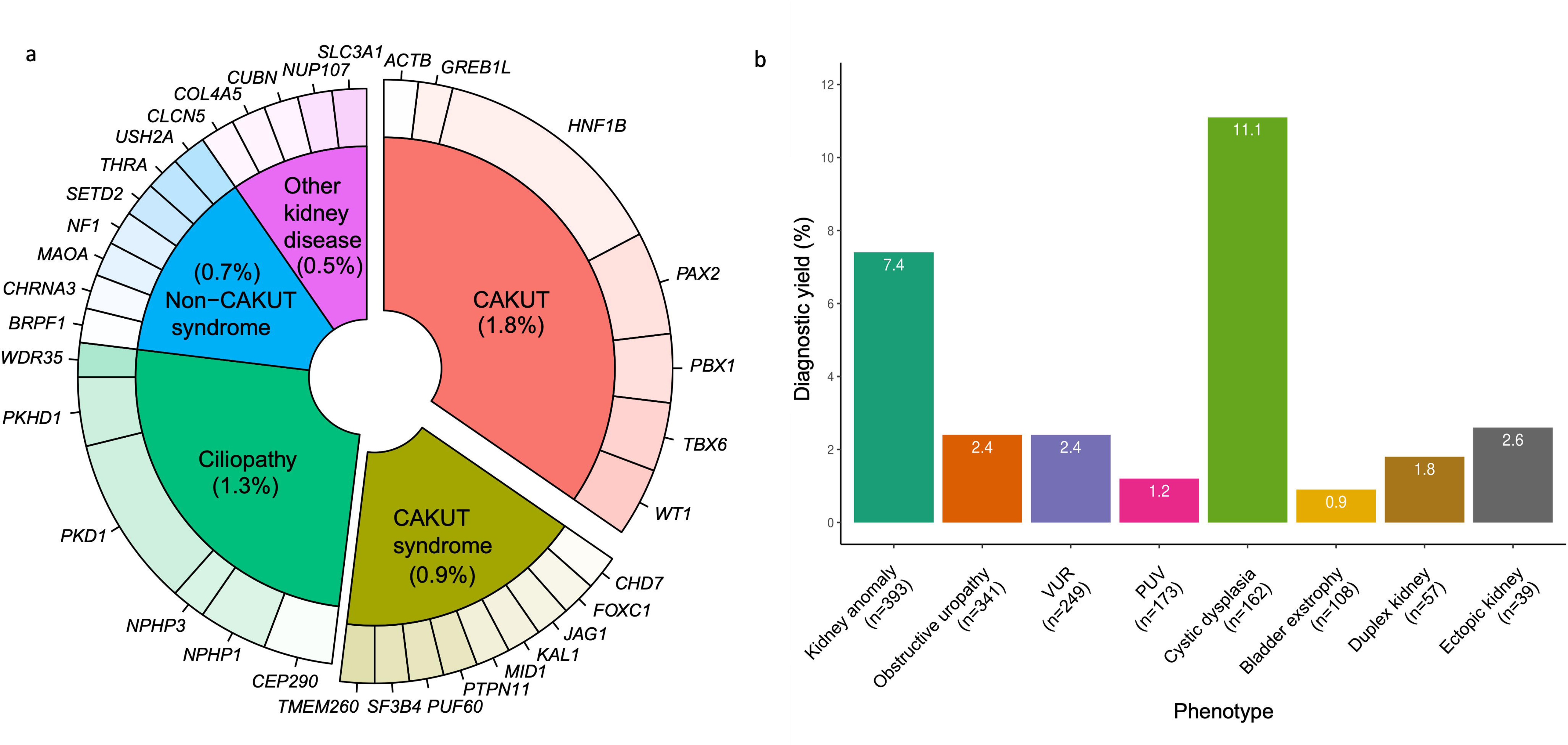
Genomic testing in 1,052 patients with CAKUT. a) Disease-causing genes identified in 4.9% of the cohort. Percentages relate to the entire cohort of 1,052 patients. b) Diagnostic yield by phenotype. CAKUT, congenital anomalies of the kidneys and urinary tract; VUR, vesico-ureteral reflux; PUV, posterior urethral valves.

As a sensitivity analysis, we recalculated the diagnostic yield excluding participants with a possible alternative diagnosis - 12 with a non-CAKUT syndrome, five with non-CAKUT diagnoses (NF1, CLCN5, COL4A5, CUBN and SLC3A1) and 12 with cystic/ciliopathy disease (PKD1, PKHD1, NPHP1, NPHP3, CEP290) - reducing the yield to 3.1% (23/743). This approach, however, cannot exclude the possibility of broader phenotypic misclassification within the remaining cohort. Overall, these data demonstrate that the prevalence of CAKUT-associated monogenic disease, at least in this cohort, is lower than previously reported and confirm that CAKUT is both genetically and phenotypically heterogeneous.

### Predictors of a genomic diagnosis

Individuals with cystic dysplasia and other kidney anomalies (i.e. kidney agenesis, hypoplasia, or dysplasia) were more likely to receive a genomic diagnosis (11.1% and 7.4%, respectively; Figure 1b). Pathogenic variants affecting HNF1B (including 17q12 deletions) accounted for 27.8% (5/18) of genomic diagnoses in the cystic dysplasia cohort, however most patients (55.6%) were found to have phenocopies; autosomal dominant polycystic kidney disease (ADPKD), autosomal recessive polycystic kidney disease (ARPKD), or nephronophthisis. Importantly, all patients with cystic dysplasia who had a confirmed genomic diagnosis also reported extra-renal features such as neurodevelopmental or skeletal abnormalities, abnormal liver function tests or hyperuricaemia (Supplementary Table S3). Likewise, in the kidney agenesis/hypodysplasia cohort 76.5% (13/17) of genomic diagnoses were associated with a syndrome or extra-renal manifestations.

Consistent with previous reports from individuals with suspected monogenic kidney disease,^18,47–49^ multivariable logistic regression analysis confirmed extra-renal features (*P*=1.1×10^-4^; OR 3.1; 95% CI 1.7-5.7), consanguinity (*P*=0.01; OR 3.0; 95% CI 1.2-6.9) and family history (*P*=0.02; OR 2.2; 95% CI 1.1-4.4) as independent predictors of a genomic diagnosis (Supplementary Table S4). A diagnosis was identified in 9.3%, 12.5% and 9.1% of those with extra-renal features, consanguinity and a family history, respectively, compared to 5%, 4.5% and 4.3% without. These findings can help clinicians identify who might benefit most from genomic testing, particularly relevant to resource-limited settings.

### Candidate gene discovery

Given known monogenic disease accounted for only a minority of cases in this cohort, an unbiased whole-exome approach was employed to identify candidate genes which may act as non-Mendelian risk factors for disease in a subset of 813 cases and 25,205 ancestry-matched controls. Single variant association testing can be underpowered when variants are rare and a collapsing strategy which aggregates variants by gene can be adopted to boost power. We therefore combined a) rare (gnomAD MAF < 0.01%) LoF (protein-truncating or splice-site) and b) rare LoF and predicted deleterious missense (REVEL ≥ 0.75) variants across 18,353 protein-coding genes. A MAF of 0.01% was chosen to capture incompletely penetrant variants and avoid loss of power associated with using an ultra-rare threshold. We did not identify any genes that were significantly enriched for rare variation after correction for multiple testing (Figure 2a; Supplementary Figure S4a; Supplementary Table S5). The median number of variants tested per gene was 5 (interquartile range [IQR] 7) and 10 (IQR 13), in each analysis respectively. The absence of exome-wide significant gene-based enrichment is likely the result of reduced power due to the genetic and phenotypic heterogeneity of the cohort and the need for multiple testing adjustment.

**Figure 2.**
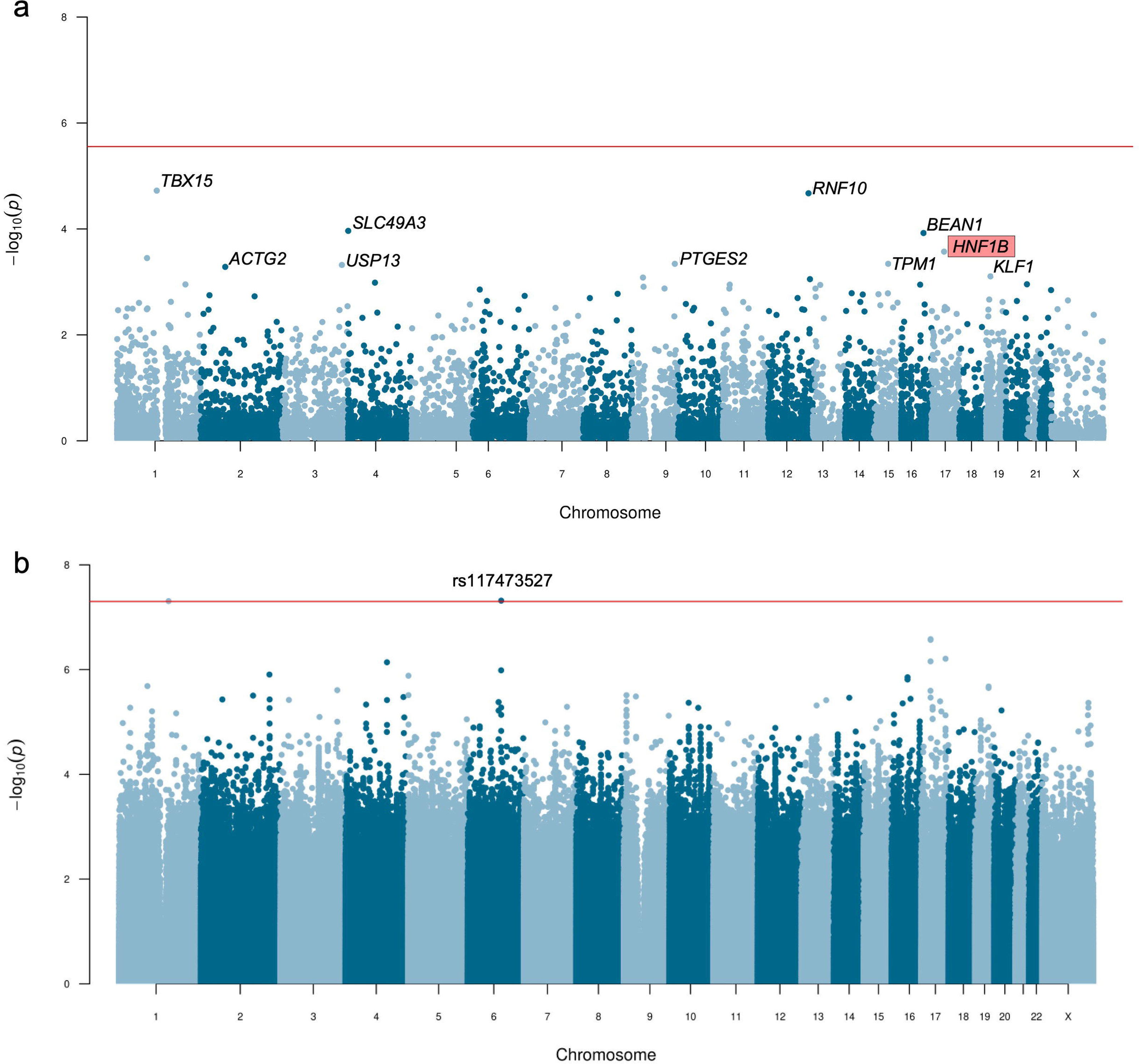
Rare and common variant association testing in 813 CAKUT cases and 25,205 ancestry-matched controls. a) Manhattan plot of gene-based rare variant association testing using SAIGE-GENE+. Qualifying variants were loss-of-function and likely deleterious missense variants with minor allele frequency < 0.01%. Each dot represents a gene. The red line indicates the exome-wide significance threshold of P=2.58×10^-6^. Genes with P < 10^-3^ are labelled and known CAKUT-gene associations are highlighted in red. b) Manhattan plot of single variant association testing using SAIGE across 10.1 million variants with MAF ≥ 0.5%. Each dot represents a variant. The red line indicates the genome-wide significance threshold of P=5×10^-8^.

The mesodermal transcription factor TBX15 showed the greatest enrichment of rare likely deleterious variation (case minor allele count [MAC] 6, control MAC 18; P = 1.9×10^-5^) and is closely related to known CAKUT gene TBX18. ^50^ HNF1B demonstrated non-significant enrichment of rare, LoF and missense variation (case MAC 6, control MAC 25; P = 2.7×10^-4^) but not LoF-only variation (case MAC 1, control MAC 2; P = 0.07) suggesting that LoF variants are not the primary drivers of the observed signal in this cohort. Excluding 33 individuals with a known monogenic diagnosis and depleting for PUV and bladder exstrophy cases did not reveal any additional signals in the remaining 539 unsolved cases. The enrichment previously seen in HNF1B was attenuated (case MAC 3, control MAC 27; P = 0.01).

To increase power, the rare variant analysis was repeated stratifying by phenotype. No statistically significant associations for a) LoF only or b) LoF and likely deleterious missense variant sets were detected (Supplementary Tables S6-S11). In the cystic dysplasia cohort (n=112), greatest enrichment was seen in PKD1 (case MAC 4, control MAC 41; P = 1.5×10^-5^), while PKD2 and HNF1B were non-significant (P = 0.85 and P = 0.72, respectively). In the congenital obstructive uropathy cohort (n=177), no known monogenic causes of functional or anatomical lower urinary tract obstruction were enriched for rare coding variation. Supplementary Tables S6-11 detail per-gene results for each subgroup: kidney anomalies (n=237), obstructive uropathy (n=177), VUR (n=174), PUV (n=132), cystic dysplasia (n=112) and bladder exstrophy (n=97).

To ensure test statistics were not inflated by the inclusion of individuals from different ancestries, we repeated the rare variant analysis using a subset of 611 cases and 19,576 controls of European ancestry (Supplementary Table S12). Both the lead gene in the mixed-ancestry analysis, *TBX15* (case MAC 3, control MAC 14; *P* = 4.2×10^-3^) and *HNF1B* (case MAC 5, control MAC 18; *P* = 3.7×10^-4^) had an attenuated signal, likely due to reduced power. Effect estimates (Spearman’s rho=0.9) and *P* values (Spearman’s rho=0.7) were consistent across both cohorts indicating that population structure is adequately controlled in this mixed-ancestry cohort (Supplementary Figure S4b and S4c).

### seqGWAS

In the absence of significant rare variant enrichment, we next explored whether common and low-frequency variation might contribute to the genomic architecture of CAKUT, performing a sequencing-based GWAS (seqGWAS) in 813 cases and 25,205 ancestry-matched controls testing for association at 10,147,641 SNVs/indels with MAF ≥ 0.5% (Figure 2b). Lambda (λ) was 1 indicating minimal evidence of confounding by population stratification (Supplementary Figure S5a). Two variants reached genome-wide significance. The first, rs117473527 (chr6:102155812:G>C) at 6q16.3 (*P* = 4.83×10^-8^; OR 3.13; 95% CI 2.08-4.72; MAF 0.01) was seen in 3.8% of cases and 1.8% control s (Supplementary Figure S5b). rs117473527 is an intergenic variant downstream of *GRIK2* (glutamate ionotropic receptor kainite type subunit 2) which has been associated with neurodevelopmental disorders and urinary tract cancers.^51,52^ *Grik2* is expressed in the murine ureteric bud at E10.5-11.5 (Supplementary Figure S5c) when it invades the metanephric mesenchyme and *GRIK2* displays robust expression in bulk-RNAseq data from 8-10 week fetal kidneys^53^ indicating it may be important during nephrogenesis. Of note, its family member *GRIK3* is upregulated in dysplastic tubules of *HNF1B* mutant kidneys.^53^ Replication of this signal in an independent cohort is needed for validation.

The second genome-wide significant variant (1 base pair deletion at 1q21.3; rs35251516; *P* = 4.93×10^-8^; MAF 0.03) was isolated, found in a low-complexity region and likely represents a sequencing artefact. Testing for association in a European-only cohort of 623 cases and 20,060 controls across 11,303,429 variants did not identify any additional signals (Supplementary Figure S6a and S6b; λ 0.998). Significant correlation between *P*-values and effect sizes in the mixed-ancestry and European-only cohorts confirms that population stratification was well-controlled (Supplementary Figure S6c and S6d).

The absence of additional significant associations is likely the result of limited power related to the phenotypic heterogeneity of the cohort and sample size. With 813 cases (assuming a prevalence of 0.2%) we have 80% power to detect association of variants with MAF > 5% and OR > 1.8 or MAF > 1% and OR > 3 (Supplementary Figure S7). To increase power, we aggregated variants across 19,118 autosomal protein-coding genes and 50 hallmark gene-sets curated by MSigDB but did not identify any enrichment in specific genes or biological pathways (Supplementary Table S13).

seqGWAS of kidney anomaly, cystic renal dysplasia, obstructive uropathy and VUR cohorts did not identify statistically significant associations (Supplementary Material) but were underpowered to detect loci other than those with large effect sizes (e.g., OR > 4 for variants with MAF 5%). To increase power, a meta-analysis of the obstructive uropathy cohort was carried out with GWAS summary statistics from 550 congenital obstructive uropathy cases and 410,449 controls from FinnGen^37^ for a total of 727 cases and 434,900 controls across 11,564,803 variants, but no genome-wide significant variants were identified (Supplementary Figure S8). As reported previously,^19^ statistically significant association was detected at two loci in 132 individuals with PUV when compared to 23,727 ancestry-matched controls across 19,651,224 SNVs/indels: 12q24.21 near TBX5 (rs10774740; P=7.81×10^-12^; OR 0.40; 95% CI 0.31-0.52; MAF 0.37) and 6p21.1 in PTK7 (rs144171242; P=2.02×10^-8^; OR 7.20; 95% CI 4.08-12.70; MAF 0.007). Both associations were replicated in an independent European cohort of 395 PUV patients (333 German and Polish patients from the CaRE for LUTO study and 62 from the UK) and 4,151 unaffected male 100KGP controls. ^19^

### Bladder Exstrophy seqGWAS

Classic bladder exstrophy (CBE) is the most frequent form of the bladder-exstrophy-epispadias-complex (BEEC) spectrum resulting from a defect in abdominal midline development. It is usually sporadic, not generally associated with other malformations and its pathogenesis is poorly understood. Using the seqGWAS approach we tested 10,103,261 SNV/indels with MAF ≥ 0.5% for association in 97 individuals with CBE and 25,921 ancestry-matched controls (Supplementary Figure S9a and S9d). Suggestive evidence of association was identified at 5q35.3 (rs147504710; *P* = 1.11×10^-7^; Supplementary Figure S9b) and 20p11.22 (P = 2.13×10^-7^; Supplementary Figure S9c; Supplementary Table S14). Of note, these variants had MAF ∼1% and a large effect size meaning they may have been missed using conventional genotyping and imputation. rs6106456 at 20p11.22 did not replicate in an independent cohort of 84 unrelated European patients and 10,804 controls (Cochran-Armitage trend test P = 0.52), however the estimated power (0.67) was insufficient to definitively confirm or refute the association. Meta-analysis with a previously published European GWAS of 628 patients and 7,352 controls^21^ across 4,663,819 variants with MAF ≥ 1% failed to identify any additional novel loci (Supplementary Figure S10).

The previously reported ISL1 association at 5q11.1 was replicated in our study with the lead variant rs9291768 (chr5:51421959:C>T) achieving a P value of 1.31×10^-3^ (OR 1.63; 95% CI 1.21-2.19; MAF 0.35). To determine whether there were any additional rare variants that might be driving this signal we repeated association testing at this locus using variants with MAC > 3 (Supplementary Figure S9e). A rare, intergenic indel rs550737686 (chr5:51494837:CCT>C) demonstrated strongest association (P = 2.35×10^-5^; OR 6.11; 95% CI 3.11-12.03; MAF 0.008). Bayesian fine mapping using functional annotations including conservation, transcription factor binding and cis-regulatory elements found rs115201978 (chr5:51035061:G>A) to be likely causal with a high posterior probability > 0.99. rs115201978 is a rare, intergenic variant (MAF 0.006) with a high CADD score of 16.9 reaching a P-value of 1.28×10^-4^ (OR 6.26; 95% CI 2.92-13.44). It is not in linkage disequilibrium (LD) with the lead indel rs550737686 (r^2^=0.1) which was excluded from the fine-mapping analysis as it is absent from the 1000 Genomes data.

### Heritability and polygenic risk

The proportion of trait heritability attributable to low-frequency and common variation can be evaluated using either WGS data or GWAS summary statistics. In a subset of 623 European CAKUT cases and 20,060 ancestry-matched controls, we used WGS data to estimate the proportion of phenotypic variance attributed to additive common and low-frequency variation (MAF ≥ 0.1%) was 23% (95% CI 1-45%); Supplementary Table S15), although this was associated with substantial uncertainty due to the limited sample size. Variants with MAF between 1% and 5% and low LD scores, meaning they are not usually inherited together with other nearby variants and therefore potentially under negative selection, accounted for >75% of this estimated heritability (Figure 3a). Heritability estimates from two additional tools (using GWAS summary statistics as opposed to WGS data) across ∼1m HapMap3 variants (MAF ≥ 1%) demonstrated broadly consistent results although with imprecise, overlapping confidence intervals, reflecting differences in the underlying models (GCTA-GREML, 10% (95% CI 0-21%); SumHer 23% (95% CI 0-54%); LDSC 18% (95% CI 0-55%).

**Figure 3.**
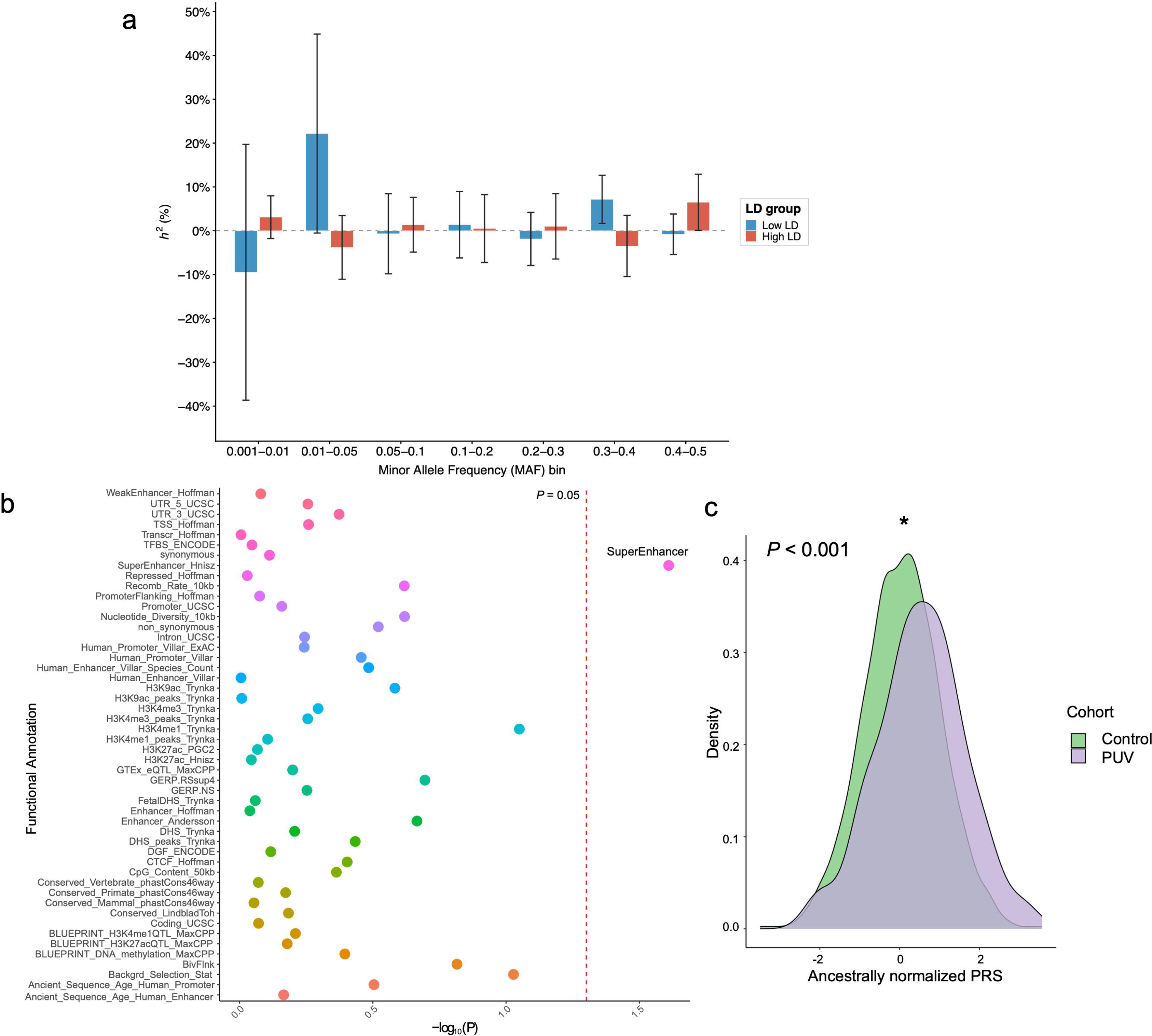
Common and low-frequency variants may explain some of the phenotypic variance in CAKUT. a) Partitioning of estimated heritability by minor allele frequency (MAF) and linkage disequilibrium in 623 CAKUT cases and 20,060 controls of European ancestry. Narrow-sense heritability (h) is represented using the liability threshold model based on a population disease prevalence of 0.2%. Error bars represent 95% confidence interval s. Negative lower confidence bounds are an expected consequence of unconstrained GREML estimation and indicate statistical uncertainty rather than biologically negative heritability. b) Enrichment of common variant heritability (MAF > 5%) across functional annotations estimated using the European CAKUT seqGWAS summary statistics. The dashed grey line represents nominal P-value of 0.05 but no annotations were significantly enriched using a false discovery rate of 10%. Super-enhancers are large clusters of transcriptional enhancers that drive expression of genes that define cell identity from Hsinz et al. c) Density plot with the distribution of the European PUV-GRS in an independent cohort of 77 cases (AGORA_PUV) and 2,746 controls. *P<0.001. PUV, posterior urethral valves; PRS, polygenic risk score.

Partitioning this heritability across functional annotations using LDSC^44^ showed nominal (*P* < 0.05) enrichment of ‘super enhancers’ – large clusters of transcriptional enhancers that drive expression of genes – but this was not significant at a false discovery rate of 10% (Figure 3b). Given the uncertainty around these estimates, they should be interpreted cautiously and as compatible with, but not conclusive evidence for, a contribution of common variation to CAKUT susceptibility.

Finally, to test whether there was a polygenic contribution to PUV we generated a PUV-GRS using 675,086 autosomal variants from the European-only male PUV seqGWAS^19^ (MAF > 0.5%; 89 cases and 8,303 controls) and applied this to WGS data from 77 individuals with PUV from the AGORA biobank and 2,746 European male controls from the 100KGP (Figure 3c; Supplementary Table S16). The ancestrally normalised PUV-GRS was significantly elevated in this independent European cohort compared to controls (P < 0.001; AUC 0.63 (95% CI 0.56-0.70); Supplementary Figure S11). The proportion of phenotypic variance estimated to be explained by the PUV-GRS had a wide confidence interval reflecting the uncertainty associated with the small sample size (liability-scale R^2^ 0.17 (95% CI 0.01-0.66)).

## Discussion

In this large cohort of patients with CAKUT, virtual panel interpretation of clinical-grade WGS identified a monogenic cause in 4.9%. The highest diagnostic yield was seen in those with kidney agenesis/hypodysplasia, cystic dysplasia, a family history, consanguinity and/or extra-renal features. This figure is at the lower end of estimates from previous studies using targeted gene panels or exome sequencing which report diagnostic yields up to 27%.^11,12,14,16,17,48,49,55–59^ This is likely related to the broad definition of phenotypes recruited to 100KGP, including individuals with mild disease, and despite the higher diagnostic yield expected with WGS over exome sequencing.^60^ WGS provides a comprehensive assessment of all variant types (SNV, indel, SVs) across the coding and non-coding genome and is less susceptible to sequencing bias.^27,28^ It is also possible individuals with a clearly monogenic cause of CAKUT were not recruited to the 100KGP, although in a cohort similar to this one, a panel of 208 genes identified a diagnosis in only 1.3% of 453 individuals.^10^ Overall, these data suggest that a minority of CAKUT patients in general nephrology practice have a monogenic cause for their condition identified and that genomic testing should be targeted towards those with kidney anomalies (particularly cystic dysplasia), a family history, consanguinity and/or extra-renal manifestations.

Unbiased interrogation of rare variation across the exome did not identify any significant gene-based enrichment. This lack of observed enrichment may be the result of several factors which limit power for discovery; a) the cohort is phenotypically heterogenous, b) this is a genetically heterogenous disorder with over 50 monogenic causes described, c) the sample size is small for a rare variant association study, and d) use of a stringent Bonferroni adjustment for multiple testing.

We used a sequencing-based GWAS approach to interrogate common and low-frequency variants. While statistically significant association was detected at 6q16.3 downstream of GRIK2, further replication in an independent cohort is necessary to confirm the association. While the GWAS itself identified limited locus-level signal, the heritability estimates suggest that common genomic variation may contribute to disease risk and that common and low-frequency variants may be enriched in cis-regulatory elements (such as enhancers), potentially impacting transcriptional networks important for kidney and urinary tract development. However, these heritability estimates are imprecise because of the modest sample size and should be interpreted as hypothesis-generating rather than definitive. We also observed differences in the heritability estimates depending on the approach used with large 95% confidence interval s, likely reflecting differences in the use of individual-level versus summary-statistic data and different modelling of LD and MAF-dependent architecture.

Previous work has shown susceptibility to lower urinary tract phenotypes, including PUV,^19^ bladder exstrophy,^21,61,62^ and VUR,^20,63,64^ is influenced by non-coding common variation. This is reinforced by the generation and validation of a low-frequency and common variant PUV-GRS. Larger, better-powered cohorts will be essential to identify these variants with small effect size and the genes/pathways they impact to generate insights into the molecular mechanisms underlying these malformations.

The main strength of this analysis lies in the use of WGS data which enables ancestry-independent variant detection across the allele frequency spectrum which is not possible using conventional genotyping arrays. Using cases and controls sequenced on the same platform minimizes the risk of confounding due to sequencing artefacts or differences in variant calling. The analysis also includes individuals of all ancestries, using a robust approach to minimize confounding by population stratification.

We acknowledge several limitations to this work. First, the molecular diagnoses are largely reliant on the Genomics England pipeline.^65^ Second, the analysis is dependent on accurate phenotyping by the recruiting team and patient’s clinician. This is highlighted by the inclusion of a few patients with other kidney phenotypes/phenocopies in the CAKUT cohort. Third, we are unable to quantify the number of patients who were not recruited to the 100KGP because a genetic diagnosis had already been made. While this means those with syndromic or clearly monogenic disease may have been excluded, genetic testing for CAKUT was not widely implemented in the UK prior to the 100KGP. Lastly, is the issue of power. In a cohort of this size, we had 80% power to identify common variants (MAF > 1%) with an OR > 3, but weaker effects or genes accounting for a small proportion of cases are not expected to be reliably detected.

In summary, we provide support for the hypothesis that non-Mendelian genomic factors play an important role in the pathogenesis of CAKUT as evidenced by the lack of an identifiable monogenic cause in most cases and the potential polygenic basis suggested by heritability estimates and GWAS. Larger cohorts are now needed to refine these heritability estimates, identify additional genomic susceptibility loci and investigate the mechanisms by which they might be impacting cell-type specific gene-regulatory networks important for urinary tract development.

## Data Sharing Statement

Supplementary material is available online at www.kidney-international.org. Summary statistics can be found at https://doi.org/10.5281/zenodo.18040141. Genomic and phenotype data from participants recruited to the 100,000 Genomes Project can be accessed by application to Genomics England Ltd at https://www.genomicsengland.co.uk/join-us/. Details of the WGS aggregated dataset used for the analysis can be found at https://re-docs.genomicsengland.co.uk/aggv2/. Code for the case–control ancestry-matching algorithm can be found at https://github.com/APLevine/PCA_Matching. Details of the rare-variant workflow can be found at https://re-docs.genomicsengland.co.uk/avt/. Details of the common-variant GWAS workflow can be found at https://re-docs.genomicsengland.co.uk/gwas/. CAKUT GWAS summary statistics will be made publicly available via the GWAS Catalog on publication (https://www.ebi.ac.uk/gwas/).

## Data Availability

Summary statistics can be found at https://doi.org/10.5281/zenodo.18040141. Genomic and phenotype data from participants recruited to the 100,000 Genomes Project can be accessed by application to Genomics England Ltd at https://www.genomicsengland.co.uk. Details of the WGS aggregated dataset used for the analysis can be found at https://re-docs.genomicsengland.co.uk/aggv2/. Code for the case control ancestry matching algorithm can be found at https://github.com/APLevine/PCA_Matching. Details of the rare variant workflow can be found at https://re-docs.genomicsengland.co.uk/avt/. Details of the common variant GWAS workflow can be found at https://re-docs.genomicsengland.co.uk/gwas/. CAKUT GWAS summary statistics will be made publicly available via the GWAS Catalog on publication (https://www.ebi.ac.uk/gwas/).

https://www.genomicsengland.co.uk/about-gecip/joining-research-community/

## Acknowledgements

The authors gratefully acknowledge the participation of the patients, and their families recruited to the 100KGP. Data from the National Genomic Research Library (NGRL) used in this research are available within the secure Genomics England Research Environment. Access to NGRL data is restricted to adhere to consent requirements and protect participant privacy. Access to NGRL data is provided to approved researchers who are members of the Genomics England Research Network, subject to institutional access agreements and research project approval under participant-led governance. For more information on data access, visit: https://www.genomicsengland.co.uk/research.

## Author contributions

DPG and MMYC conceived the study and designed the experiments. MMYC, OSA and SP analysed the data and interpreted the results. APL, DB and HCS provided expertise in genomic analysis and contributed to study design. CDV provided computational support. Clinical phenotyping and genotyping/sequencing was carried out by the remaining co-authors (AGORA, ArtDECO Consortium, CaRE for LUTO, bladder exstrophy cohort). The manuscript was drafted by MMYC and critically revised by all authors. All authors approved the final manuscript and agree to be accountable for all aspects of the work.

## Supplementary Material

**Supplementary Methods**

**Supplementary Figure S1.** Study workflow.

**Supplementary Figure S2.** Ancestry matching of cases and controls.

**Supplementary Figure S3.** Human Phenotype Ontology (HPO) terms in the CAKUT cohort.

**Supplementary Figure S4.** Quantile-quantile plot of CAKUT rare variant gene-based analyses.

**Supplementary Figure S5.** Quantile-quantile and regional association plot for CAKUT seqGWAS.

**Supplementary Figure S6.** European-only CAKUT seqGWAS.

**Supplementary Figure S7.** Statistical power for CAKUT GWAS.

**Supplementary Figure S8.** Meta-analysis of 727 cases and 434,900 controls with congenital obstructive uropathy.

**Supplementary Figure S9.** seqGWAS of 97 bladder exstrophy cases and 22,037 ancestry-matched controls.

**Supplementary Figure S10.** Meta-analysis of 725 cases and 29,389 controls with bladder exstrophy.

**Supplementary Table S1.** List of human phenotype ontology (HPO) codes used to stratify CAKUT cases.

**Supplementary Table S2.** List of SNOMED-CT and ICD-10 codes used to filter out controls with known kidney disease.

**Supplementary Table S3.** Genetic and phenotypic data for CAKUT cases with a genetic diagnosis.

**Supplementary Table S4.** Predictors of a genetic diagnosis in CAKUT.

**Supplementary Table S5.** Summary statistics from the rare variant gene-based analysis of 813 cases and 25,205 ancestry-matched controls.

**Supplementary Table S6.** Summary statistics from the rare variant gene-based analysis of 237 kidney anomaly cases and 22,733 ancestry-matched controls.

**Supplementary Table S7.** Summary statistics from the rare variant gene-based analysis of 177 congenital obstructive uropathy cases and 24,451 ancestry-matched controls.

**Supplementary Table S8.** Summary statistics from the rare variant gene-based analysis of 112 cystic dysplasia cases and 24,084 ancestry-matched controls.

**Supplementary Table S9.** Summary statistics from the rare variant gene-based analysis of 174 VUR cases and 22,562 ancestry-matched controls.

**Supplementary Table S10.** Summary statistics from the rare variant gene-based analysis of 132 PUV cases and 23,727 ancestry-matched controls.

**Supplementary Table S11.** Summary statistics from the rare variant gene-based analysis of 97 bladder exstrophy cases and 22,037 ancestry-matched controls.

**Supplementary Table S12.** Summary statistics from the rare variant gene-based analysis of 611 European CAKUT cases and 19,576 ancestry-matched controls.

**Supplementary Table S13.** Pathway analysis for 813 CAKUT cases versus 25,205 ancestry-matched controls.

**Supplementary Table S14.** Lead variants from bladder exstrophy seqGWAS.

**Supplementary Table S15.** Heritability estimation in 623 European CAKUT cases and 20,060 controls using GREML-LDMS.

**Supplementary Table S16.** Variants and weights used for PUV Genomic Risk Score.

**Supplementary References**

