## Supplementary Material for "Whole-genome sequencing characterizes monogenic and polygenic contributions to structural kidney and urinary tract malformations"

**Supplementary Methods.**

**Supplementary Figure S1.** Study workflow.

**Supplementary Figure S2.** Ancestry matching of cases to controls.

**Supplementary Figure S3.** Human Phenotype Ontology (HPO) terms in the CAKUT cohort.

**Supplementary Figure S4.** Quantile-quantile plot of CAKUT rare variant gene-based analyses.

**Supplementary Figure S5.** Quantile-quantile and regional association plot for CAKUT seqGWAS.

**Supplementary Figure S6.** European-only CAKUT seqGWAS.

**Supplementary Figure S7.** Statistical power for CAKUT GWAS.

**Supplementary Figure S8.** Meta-analysis of 727 cases and 434,900 controls with congenital obstructive uropathy.

**Supplementary Figure S9.** seqGWAS of 97 bladder exstrophy cases and 22,037 ancestry-matched controls.

**Supplementary Figure S10.** Meta-analysis of 725 cases and 29,389 controls with bladder exstrophy.

**Supplementary Figure S11.** Receiver Operating Characteristic (ROC) curve for PUV-GRS.

**Supplementary Table S1.** List of human phenotype ontology (HPO) codes used to stratify CAKUT cases.

**Supplementary Table S2.** List of SNOMED-CT and ICD-10 codes used to filter out controls with known kidney disease.

**Supplementary Table S3.** Genetic and phenotypic data for CAKUT cases with a genetic diagnosis.

**Supplementary Table S4.** Predictors of a genetic diagnosis in CAKUT.

**Supplementary Table S5.** Summary statistics from the rare variant gene-based analysis of 813 cases and 25,205 ancestry-matched controls.

**Supplementary Table S6.** Summary statistics from the rare variant gene-based analysis of 237 kidney anomaly cases and 22,733 ancestry-matched controls.

**Supplementary Table S7.** Summary statistics from the rare variant gene-based analysis of 177 congenital obstructive uropathy cases and 24,451 ancestry-matched controls.

**Supplementary Table S8.** Summary statistics from the rare variant gene-based analysis of 112 cystic dysplasia cases and 24,084 ancestry-matched controls.

**Supplementary Table S9.** Summary statistics from the rare variant gene-based analysis of 174 VUR cases and 22,562 ancestry-matched controls.

**Supplementary Table S10.** Summary statistics from the rare variant gene-based analysis of 132 PUV cases and 23,727 ancestry-matched controls.

**Supplementary Table S11.** Summary statistics from the rare variant gene-based analysis of 97 bladder exstrophy cases and 22,037 ancestry-matched controls.

**Supplementary Table S12.** Summary statistics from the rare variant gene-based analysis of 611 European CAKUT cases and 19,576 ancestry-matched controls.

**Supplementary Table S13.** Pathway analysis for 813 CAKUT cases versus 25,205 ancestry-matched controls.

**Supplementary Table S14.** Lead variants from bladder exstrophy seqGWAS.

**Supplementary Table S15.** Heritability estimation in 623 European CAKUT cases and 20,060 controls using GREML-LDMS.

**Supplementary Table S16.** Variants and weights used for PUV Genomic Risk Score.

**Supplementary References**

**Supplementary Methods**

### The 100,000 Genomes Project

The 100,000 Genomes Project (100KGP) dataset (version 19) consists of WGS data, clinical phenotypes encoded using Human Phenotype Ontology (HPO) codes, and retrospective and prospectively ascertained hospital records for 90,173 individuals with cancer or rare disease, as well as their unaffected relatives.^S1^ Version 19 data release was used for the diagnostic yield analysis. The Aggregated Variant Call (AggV2; release 10) dataset consisting of 78,195 germline genomes aligned to GRCh38 was used for the association analyses. Ethical approval for the 100KGP was granted by the Research Ethics Committee for East of England – Cambridge South (REC Ref 14/EE/1112). The study workflow is shown in Supplementary Figure S1.

### DNA preparation and extraction

99% of DNA samples were extracted from blood and prepared using EDTA, with the remaining 1% sourced from saliva, tissue and fibroblasts. Samples underwent quality control assessment based on concentration, volume, purity and degradation. Libraries were prepared using the Illumina TruSeq DNA PCR-Free High Throughput Sample Preparation kit or the Illumina TruSeq Nano High Throughput Sample Preparation kit.

### Whole-genome sequencing, alignment and variant calling

Samples were sequenced with 150bp paired-end reads using an Illumina HiSeq X and processed on the Illumina North Star Version 4 Whole Genome Sequencing Workflow (NSV4, version 2.6.53.23), comprising the iSAAC Aligner (version 03.16.02.19) and Starling Small Variant Caller (version 2.4.7). Samples were aligned to the Homo Sapiens NCBI GRCh38 assembly. Alignments had to cover ≥ 95% of the genome at ≥ 15X with mapping quality > 10 in order for samples to be retained. Samples achieved a mean of 97.4% coverage at 15X with a median genome-wide coverage of 39X. Samples with < 2% cross contamination as determined by the VerifyBamID algorithm were kept. Copy number and structural variant (>50bp) calling was performed using CANVAS (version 1.3.1)^S2^ and MANTA (version 0.28.0)^S3^ respectively.

### gVCF annotation and variant-level quality control

We used the Genomics England Aggregated Variant Calls (AggV2) for the association analysis. AggV2 comprises 78,195 germline genomes aligned to GRCh38 from data release 10; [https://redocs.genomicsengland.co.uk/aggv2/.](https://re-docs.genomicsengland.co.uk/aggv2/) gVCFs were aggregated in batches of 1,000 using gvcfgenotyper (Illumina, version: 2019.02.26, [https://github.com/Illumina/gvcfgenotyper)](https://github.com/Illumina/gvcfgenotyper) with variants normalized, left-aligned and multi-allelic variants decomposed using vt (version 0.57721). Variants were retained if they passed the following filters: missingness ≤ 5%, median depth ≥ 10, median GQ ≥ 15, percentage of heterozygous calls not showing significant allele imbalance for reads supporting the reference and alternate alleles (ABratio) ≥ 25%, percentage of complete sites (completeGTRatio) ≥ 50% and *P* value for deviations from Hardy-Weinberg equilibrium (HWE) in unrelated samples of inferred European ancestry ≥ 1×10^-5^. Male and female subsets were analyzed separately for sex chromosome quality control. Annotation was performed using Variant Effect Predictor (VEP, version 98.2)^S4^ including CADD (version 1.5),^S5^ and allele frequencies from publicly available databases including gnomAD (version 3)^S6^ and TOPMed (Freeze 5).^S7^ Variants were filtered using bcftools (version 1.11).^S8^

### Cohort Selection

Patients were recruited to the 100KGP as part of the ‘Congenital anomalies of the kidneys and urinary tract (CAKUT)’ cohort with the following inclusion criteria: CAKUT with syndromic manifestations in other organ systems; isolated CAKUT with a first-degree relative with CAKUT or unexplained CKD; multiple distinct kidney/urinary tract anomalies; CAKUT with unexplained end-stage kidney disease before the age of 25 years. Pre-screening for *HNF1B* and *SALL1* was recommended with a personal or family history of diabetes, or imperforate anus, ear or thumb abnormalities, respectively. Testing for other CAKUT genes was not routinely undertaken. Those with a known genetic or chromosomal abnormality were excluded. Individuals with kidney cysts (not dysplasia) were recruited to the 100KGP ‘cystic kidney disease’ cohort.^S9^ Five probands had documented negative genetic testing prior to recruitment with either targeted sequencing and/or microarray-based comparative genomic hybridisation (array CGH).

For the association analyses, patients were stratified by HPO terms (Supplementary Table S1) into the following six groups: kidney anomalies (n=237), cystic renal dysplasia (n=112), obstructive uropathy (excluding posterior urethral valves; n=177), VUR (n=174), PUV (n=132) and bladder exstrophy (n=97).

Unaffected relatives of non-kidney disease participants from the 100KGP, excluding those with SNOMED-CT or ICD-10 codes consistent with kidney disease/failure (Supplementary Table S2) were used as controls. A cohort of unrelated ancestry-matched cases and controls was generated to minimize the effects of population stratification in this mixed-ancestry cohort (see below; Supplementary Figure S2).

### Relatedness estimation and principal components analysis

A set of 127,747 high quality autosomal linkage-disequilibrium (LD) pruned biallelic single nucleotide variants (SNVs) with minor allele frequency (MAF) > 1% was generated using PLINK (version 1.9).^S10^ SNVs were included if they met all of the following criteria: missingness < 1%, median GQ ≥ 30, median depth ≥ 30, AB Ratio ≥ 0.9, completeness ≥ 0.9. Ambiguous SNVs (AC or GT) and those in a region of longrange high LD (<https://genome.sph.umich.edu/wiki/Regions_of_high_linkage_disequilibrium_(LD))> were excluded. LD pruning was carried out using an r^2^ threshold of 0.1 and window of 500kb. SNVs out of HWE in any of the AFR, EAS, EUR or SAS 1000 Genomes populations were removed (pHWE < 1 ×10^-5^). Using this variant set, a pairwise kinship matrix was generated using the PLINK2^S11^ implementation of the KING-Robust algorithm^S12^ and a subset of unrelated samples was ascertained using a kinship coefficient threshold of 0.0884 (2nd degree relationships). 7 cases and 1,942 controls were found to be related and removed, leaving 817 cases and 25,718 controls. Ten principal components were generated using PLINK2^S11^ for downstream ancestry-matching and use as covariates in the association analyses.

### Ancestry-matching of cases and controls

Given the mixed-ancestry composition of the cohort we employed a case-control ancestry-matching algorithm to optimize genomic similarity and minimize the effects of population structure. A custom R script ([https://github.com/APLevine/PCA_Matching)](https://github.com/APLevine/PCA_Matching) was used to match cases to controls within a distance threshold calculated using the top ten principal components weighted by the percentage of genetic variation explained by each component (Supplementary Figure S2a). Only controls within a userdefined specified distance of a case were included with each case having to match a minimum of two controls to be included in the final cohort. A total of four cases and 513 controls were excluded using this approach, leaving 813 cases and 25,205 controls. Stratification by subphenotype resulted in the following unrelated, ancestry-matched cohorts for downstream analysis: kidney anomalies (237 cases and 25,781 controls); obstructive uropathy (177 cases and 25,841 controls); VUR (174 cases and 25,844 controls); PUV (132 cases and 10,425 controls; all male); cystic dysplasia (112 cases and 25,906 controls); bladder exstrophy (97 cases and 25,921 controls).

For European-only cohort analyses, we used the Genomics England random forest model to predict ancestries based on 1000 Genomes (Phase 3) sub-populations (https://re-

docs.genomicsengland.co.uk/ancestry_inference/). Individuals with > 0.8 probability of being of European ancestry were included in downstream analyses and compared to 20,060 European controls:

CAKUT (623 cases); kidney anomalies (194 cases); obstructive uropathy (142 cases); VUR (139 cases); PUV (89 cases and 8,303 controls; all male); cystic dysplasia (94 cases); bladder exstrophy (82 cases).

Supplementary Figure S2b shows the PCA for European-only cases and controls.

### Identification of disease-causing variants

Probands were assessed using the Genomics England clinical interpretation pipeline to determine a genomic diagnosis.^S1^ This workflow extracts rare (MAF < 1% for autosomal recessive and MAF < 0.1% for autosomal dominant inheritance), protein-truncating and missense variants and high-quality CNVs > 10kb (MAF < 0.5% (LOSS) and MAF < 1% (GAIN)) that intersect with an expert-curated panel of 60 CAKUT-associated genes ([https://panelapp.genomicsengland.co.uk/panels/234/v](https://panelapp.genomicsengland.co.uk/panels/234/)1.178). Additional gene panels were used depending on each patients’ HPO terms e.g intellectual disability, skeletal dysplasia.

In those without a molecular diagnosis, the top five ranked Exomiser^S13^ variants and all CNV calls across 286 expertly-curated kidney disease genes (green genes from Genomics England PanelApp: CAKUT v1.178 and unexplained kidney failure v12.7; https://panelapp.genomicsengland.co.uk/panels/156/) and the six most common genomic loci associated with syndromic CAKUT (1q21.1, 4p16.3, 16p11.2, 16p13.11, 17q12, 22q11.2)^S14^ were extracted and manually reviewed to identify additional variants meeting criteria for pathogenicity. Compound heterozygosity was confirmed where parental DNA was available (5/8 probands).

Multi-disciplinary review of candidate variants, considering segregation with disease within a family and mode of inheritance, was undertaken by local Genomic Medicine Centres with application of the

Association for Clinical Genomic Science (ACGS) Best Practice Guidelines for Variant Classification in Rare Disease (https://www.acgs.uk.com/media/12533/_media_12533_uk-practice-guidelines-for-variantclassification-v12-2024.pdf) to determine pathogenicity. These well-defined criteria objectively integrate variant information including population frequency, computational predictions of deleteriousness, functional domain localisation, putative mechanism of disease and previous associations with phenotypes in reputable databases to assign one of the following classifications: pathogenic, likely pathogenic, variant of uncertain significance (VUS), likely benign or benign.

### Aggregate rare coding variant analysis

Single variant association testing is underpowered when variants are rare and a collapsing approach which aggregates variants by gene can be adopted to boost power. We extracted coding SNVs and indels with MAF < 0.01% in gnomAD,^S6^ annotated with one of the following: missense, inframe insertion, inframe deletion, start loss, stop loss, stop gain, frameshift, splice donor or splice acceptor. Variants were further filtered by REVEL ^S15^ score using a threshold of ≥ 0.75 which offers a sensitivity of 0.55 and specificity of 0.97 for pathogenicity. ‘High confidence’ loss-of-function variants (stop gain, splice site, frameshift) were determined by LOFTEE.^S6^ Variants meeting the following quality control (QC) filters were retained: minor alelle count (MAC) ≤ 20, median site-wide depth in non-missing samples > 20 and median genotype quality (GQ) ≥ 30. Sample-level QC metrics for each site were set to minimum depth per sample of 10, minimum GQ per sample of 20 and ABratio *P* value > 0.001. Variants with significantly different missingness between cases and controls (*P* < 10^-5^) or > 5% missingness overall were excluded. We employed SAIGE-GENE+ (v1.3.6)^S16^ to ascertain whether rare coding variation was enriched in cases on a per-gene basis exome-wide. SAIGE-GENE+ utilizes a generalized mixed-model to correct for population stratification and cryptic relatedness and a leave-one-chromosome-out (LOCO) approach to prevent proximal contamination. Sex and the top ten principal components were included as fixed effects when fitting the null model. SAIGE-GENE+ collapses ultra-rare variants (MAC ≤ 10) to reduce data sparsity, improving variance estimation and reducing type 1 error rates seen with ultra-rare variants and unbalanced case-control ratios. It combines single-variant score statistics and their covariance estimates to perform SKAT-O^S17^ gene-based association testing, upweighting rarer variants using the beta(1,25) weights option. SKAT-O is a combination of a traditional burden and variance-component test and provides robust power when the underlying genetic architecture is unknown. A Bonferroni adjusted *P* value of 2.7×10^-6^ (0.05/18,353 genes) was used to determine the exome-wide significance threshold. Power was estimated to be 50% in a cohort of 813 cases and 25,205 controls using the SKAT package in R assuming 10% of rare variants (MAF < 0.001) in a gene are causal with a maximum odds ratio of 10 using SKAT-O across a typical gene-sized region (30kb).

### seqGWAS

Genome-wide single variant association analysis was carried out using the R package SAIGE (version

1.0.7)^S18^ which uses a generalized logistic mixed model (GLMM) to account for population stratification.

2,000 randomly selected high-quality, autosomal, bi-allelic, LD-pruned SNVs with MAF > 5% were used

to generate a genetic relationship matrix and fit the null GLMM. Sex and the top ten principal components were used as fixed effects. SNVs and indels with MAF > 0.5% and that passed the following quality control filters were retained: MAC ≥ 20, missingness < 2%, HWE *P* > 10^-6^ and differential missingness *P* > 10^-5^. A score test for association was performed for 10,147,641 variants across 813 cases and 25,205 controls (λ 1.00). Like SAIGE-GENE+,^S16^ SAIGE ^S18^ employs a saddlepoint approximation^S19^ to calibrate score test statistics and obtain more accurate *P* values than the normal distribution. seqGWAS was also performed for each stratified subphenotype using both mixed ancestry and European only cohorts (see table below). The R packages qqman^S20^ and GWASTools^S21^ were used to create Manhattan and Q-Q plots, and LocusZoom^S22^ to visualize regions of interest.

| Phenotype | Mixed ancestry variants tested | Mixed ancestry λ | European-only variants tested | European only λ |
| --- | --- | --- | --- | --- |
| CAKUT | 10,147,641 | 1 | 11,303,509 | 1 |
| Kidney anomalies | 10,117,134 | 1.01 | 11,300,392 | 1.01 |
| Obstructive uropathy | 10,108,380 | 1.03 | 11,294,998 | 1 |
| VUR | 10,110,379 | 1.01 | 11,305,374 | 0.99 |
| PUV | 11,367,695 | 1.01 | 11,318,098 | 1.04 |
| Cystic dysplasia | 10,107,621 | 1.01 | 11,300,462 | 1.03 |
| Bladder exstrophy | 10,103,261 | 1.03 | 11,299,045 | 1.05 |

### Gene-set analysis

MAGMA (version 1.09)^S23^ was used to test the joint association of all SNVs/indels with MAF > 0.1% within a particular gene-set. Variants were assigned to 17,636 protein coding genes (NCBI build 38) and competitive gene-set analysis performed for 50 hallmark gene sets from MsigDB (version 7.4).^S24^ (These hallmark gene sets are generated computationally, representing specific well-defined biological states or processes condensed from overlapping databases (BIOCARTA,^S25^ KEGG,^S26^ REACTOME,^S27^

WikiPathways,^S28^ PID,^S29^ GO^S30^). Competitive analysis tests whether the joint association of genes in a gene-set is stronger than a randomly selected set of similarly sized genes. Bonferroni correction was applied for the total number of tested gene sets (*P*=0.05/50=0.001).

### Meta-analysis

GWAS summary statistics from the Genomics England obstructive uropathy cohort (177 cases and 24,451 controls) were meta-analysed with publicly available data from FinnGen ^S31^ (Release 10) using the inverse variance weighted approach as implemented by METAL.^S32^ The FinnGen cohort consisted of 550 cases with congenital obstructive uropathy and 410,449 controls, excluding those with congenital malformations of the urinary system, and included imputed variants with a MAC > 5. A total of 11,647,818 common variants between the two datasets were meta-analysed. Lambda was 1.01 (Supplementary Figure S8b). Variants with a difference in minimum and maximum allele frequency > 0.2 suggesting allele mismatching (n=1,565) and heterogeneity *P* value < 0.01 (n=83,015) were excluded. The same approach was used to meta-analyse the bladder exstrophy cohort (97 cases and 22,037 controls) with GWAS summary statistics from Mingardo et al.^S33^ consisting of 628 European cases and 7,352 controls. 4,696,412 common variants with MAF ≥ 1% were meta-analysed with 32,593 variants excluded due to a heterogeneity *P* value < 0.01. Lambda was 0.98 (Supplementary Figure S10b). The R packages qqman^S20^ and GWASTools^S21^ were used to create Manhattan and Q-Q plots, and LocusZoom^22^ to visualize regions of interest.

### Statistical Fine-Mapping

PAINTOR (version 3.1)^S34^ was used to fine-map the 5q11.1 locus from the bladder exstrophy seqGWAS. This approach uses an empirical Bayes prior to integrate functional annotation data, LD patterns and strength of association to estimate the posterior probability (PP) of a variant being causal. Variants within a 100kb window centring on the lead variant with P < 0.05 were extracted (n=74) however the lead indel itself was excluded from the analysis as it was not present in the 1000 Genomes data used to calculate LD. Z-scores were calculated as effect size (β) divided by standard error. One causal variant was assumed per locus. LD matrices of pairwise correlation coefficients were derived using 1000 Genomes European data (Phase 3)^S35^ as a reference, excluding variants with ambiguous alleles (A/T or G/C). The locus was intersected with the following functional annotations downloaded using the UCSC Table Browser: GENCODE^S36^ (version 29) transcripts (wgEncodeGencodeBasicV29, updated (2019-02-15),

PhastCons^S37^ (phastConsElements100way, updated 2015-05-08), ENCODE^S38^ cCREs

(encodeCcreCombined, updated 2020-05-20), transcription factor binding clusters

(encRegTfbsClustered, updated 2019-05-16), DNase I hypersensitivity clusters

(wgEncodeRegDnaseClustered, updated 2019-01-08) and H1 Human embryonic stem cell Hi-C data

(h1hescInsitu from (Krietenstein et al. 2020)).^S39^

### Heritability

Narrow-sense heritability (*h^2^*) is an estimate of the proportion of phenotypic variation attributed to additive genetic effects. The analysis was performed using GREML-LDMS^S40^ from the GCTA package (version 1.93.1beta) in a subset of individuals with genetically defined European ancestry (623 cases and 20,060 controls). 22,052,734 variants with MAF ≥ 0.1% underwent the same quality control filtering as described above and were stratified into 28 bins based on MAF and LD. The REML (restricted maximum likelihood) function was used to conduct a GREML-LDMS analysis using the 28 genetic relationship matrices (GRMs), including the top four principal components as covariates. The observed heritability was transformed to an underlying continuous liability threshold model^S41^ adjusting for the ratio of cases to controls and a population prevalence of disease of 0.2%.^S42^ Results from three heritability estimation approaches (GCTA-GREML, LDAK-SumHer^S43^ and LD Score Regression) were compared across the same 1,094,435 autosomal, biallelic HapMap3 variants with MAF ≥ 1%. WGS data was used for GCTA-GREML with age and ten PCs as covariates. seqGWAS summary statistics (excluding MHC locus) lifted over to hg19 with UCSC LiftOver^S44^ were used for LDAK-SumHer and LDSC using European UK BioBank and 1000 Genomes (Phase 3) cohorts as LD reference panels, respectively.

### LD Score Regression

Stratified LD Score Regression (version 1.0.1)^S45^ was used to estimate the enrichment of heritability from European-only CAKUT seqGWAS data across functional genomic annotations using the baseline-LD model (version 2.2). This model includes a comprehensive set of coding, conserved, regulatory, and epigenomic annotations, accounting for local linkage disequilibrium and minor allele frequency– dependent architecture. Enrichment was calculated as the proportion of heritability explained by each annotation relative to the proportion of variants it contains, with significance assessed using block jackknife standard errors. Analyses were conducted using 1,173,583 HapMap3 variants (excluding the MHC locus) and European-ancestry LD scores from the 1000 Genomes Project (Phase 3). Lambda GC was

0.9986.

### PUV Genomic Risk Score

PRS-CS^S46^ (version 1.1.0) was used to generate effect weights (via Bayesian regression and continuous shrinkage priors) for autosomal variants present in both the PUV European seqGWAS summary statistics (89 cases and 8,303 controls) and either the Dutch AGORA PUV cases (n=77) or European 100KGP controls (n=2,746). Only males were included. For each variant, PRS-CS uses each variant’s BETA coefficient and corresponding standard error obtained from the GWAS to estimate an effect weight. Linkage disequilibrium was accounted for using a European reference panel from the UK BioBank. A global shrinkage parameter of 10^-2^ was used. This parameter is altered based on how polygenic a trait is expected to be, with smaller values generally providing effect weights with better predictive ability for less polygenic traits. In total, 675,086 autosomal variants received effect weights.

To calculate a genomic risk score (GRS) using these effect weights, pgsc_calc (version 2.0.0)^S47^ was used.

A reference panel containing the combined data of the 1000 Genomes Project (Phase 3)^S35^ and the Human Genome Diversity Project^S48^ was used to calculate PCA loadings required to ancestrally normalise the scores via a PCA-based method provided by pgsc_calc.^S47^ The ancestrally normalised GRS provided a bootstrapped 95% AUC confidence interval of 0.63 ± 0.07 when used as a predictor of PUV in a binomial GLM model. The bootstrapped liability-scale R^2 S49^ 95% confidence interval was calculated using the boot package in R and a disease prevalence of 1 in 4000. The ROC curve and AUC were generated using the pROC and ggplot2 R packages.

### Replication Cohorts

The AGORA (Aetiologic research into Genetic and Occupational/Environmental Risk Factors for

Anomalies in Children) biobank (part of the Dutch Radboud Biobank, [www.radboudbiobank.nl)](http://www.radboudbiobank.nl/) contains DNA samples, clinical data and parental questionnaires from children with major structural birth defects and childhood cancer.^S50^ WGS data from 77 unrelated male individuals with PUV and 2,746 male European controls with non-urinary tract cancers (from the 100KGP) were used to validate the PUV GRS.

An independent European cohort of 84 patients with bladder exstrophy (Prof Bill Newman, University of Manchester) was used to try and replicate the bladder exstrophy association at 20p11.22.

### Supplementary Figure S1

**The 100,000 Genomes Project (v19)**

n=90,173

CAKUT

n=824

Unaffected relatives with no renal disease

n=27,660

Unrelated CAKUT

n=817

Unrelated controls

n=25,718

Ancestry

-

matched CAKUT

n=813

Ancestry

-

matched controls

n=25,205

Association analysis

Rare variants

MAF < 0.01%

Common variants

MAF ≥ 0.5%

Exome

-

wide gene

-

based association

Meta

-

analysis:

Obstructive Uropathy

Bladder Exstrophy

seqGWAS

Heritability

estimation in

European cohort

1

,052 CAKUT

probands

GRCh38

Clinical

interpretation

using 286 kidney

disease gene panel

Loss of function

variants only

Loss of function +

missense (REVEL ≥

variants

0.75)

Validation of PUV

-

GRS in European

cohort

**Aggregated variant dataset (AggV2; v10)**

n=78,195

**Supplementary Figure S1. Study workflow.** CAKUT, congenital anomalies of the kidneys and urinary tract; GRCh38, Genome Reference Consortium Human Build 38; MAF, minor allele frequency; seqGWAS, whole-genome sequencing based genome-wide association study; GRS, genomic risk score; PUV, posterior urethral valves.

### Supplementary Figure S2

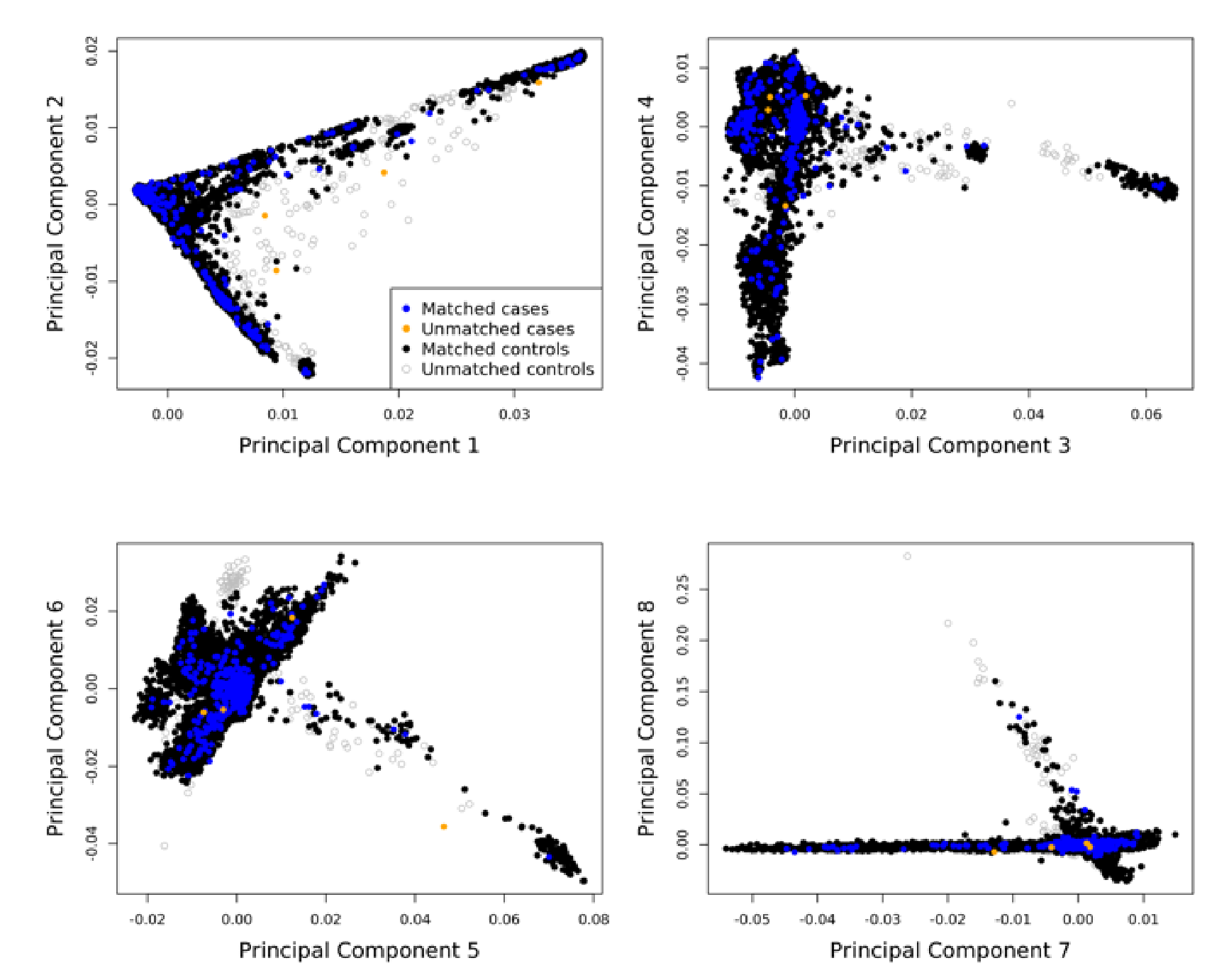

b

a

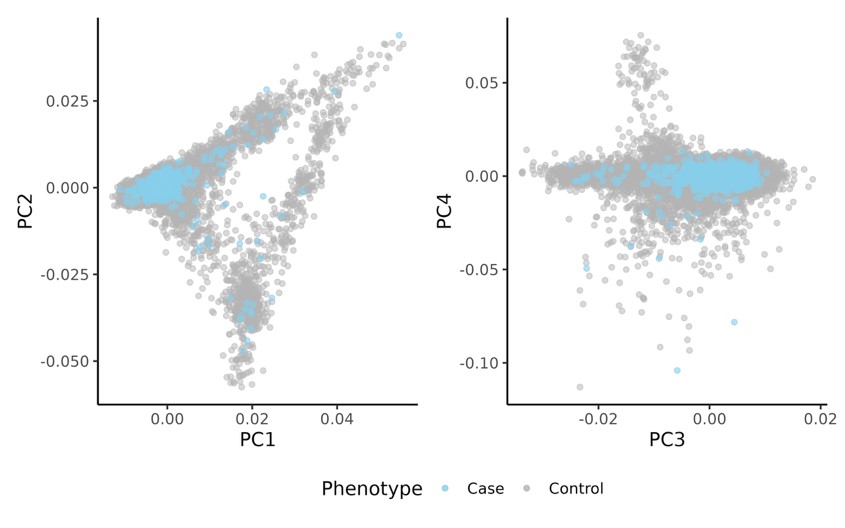

**Supplementary Figure S2. Principal component analysis showing ancestry matching of cases to controls.** a) First eight principal components for 817 ancestry-matched cases (blue) and 25,718 controls (black). Four cases (orange) and 513 (grey) controls were excluded from downstream analyses. b) First four principal components for the subset of 623 cases and 20,060 controls of European ancestry.

### Supplementary Figure S3

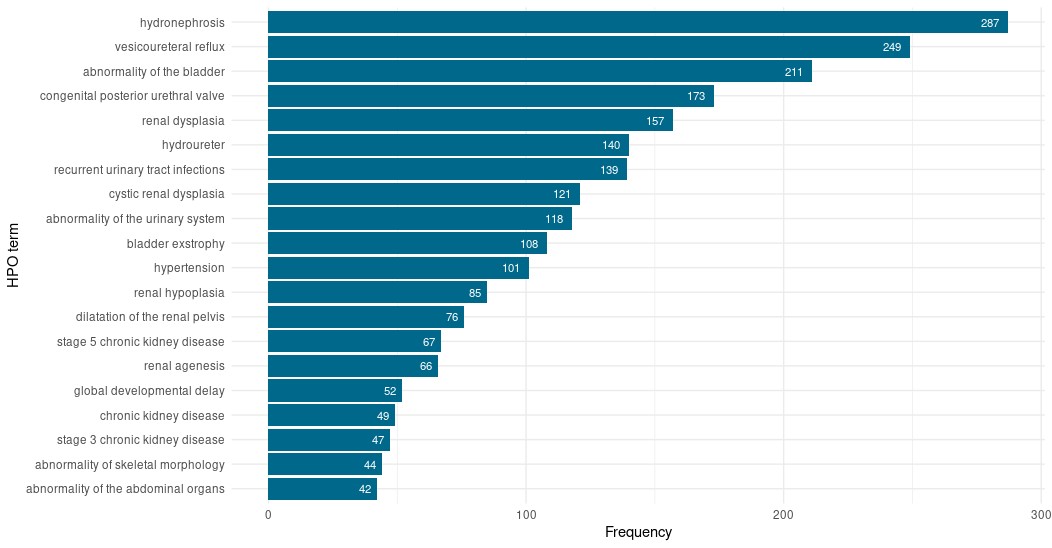

**Supplementary Figure S3.** Most frequently reported Human Phenotype Ontology (HPO) terms in the

CAKUT cohort (n=1,052). Patients could have >1 associated HPO term.

**Supplementary Figure S4**

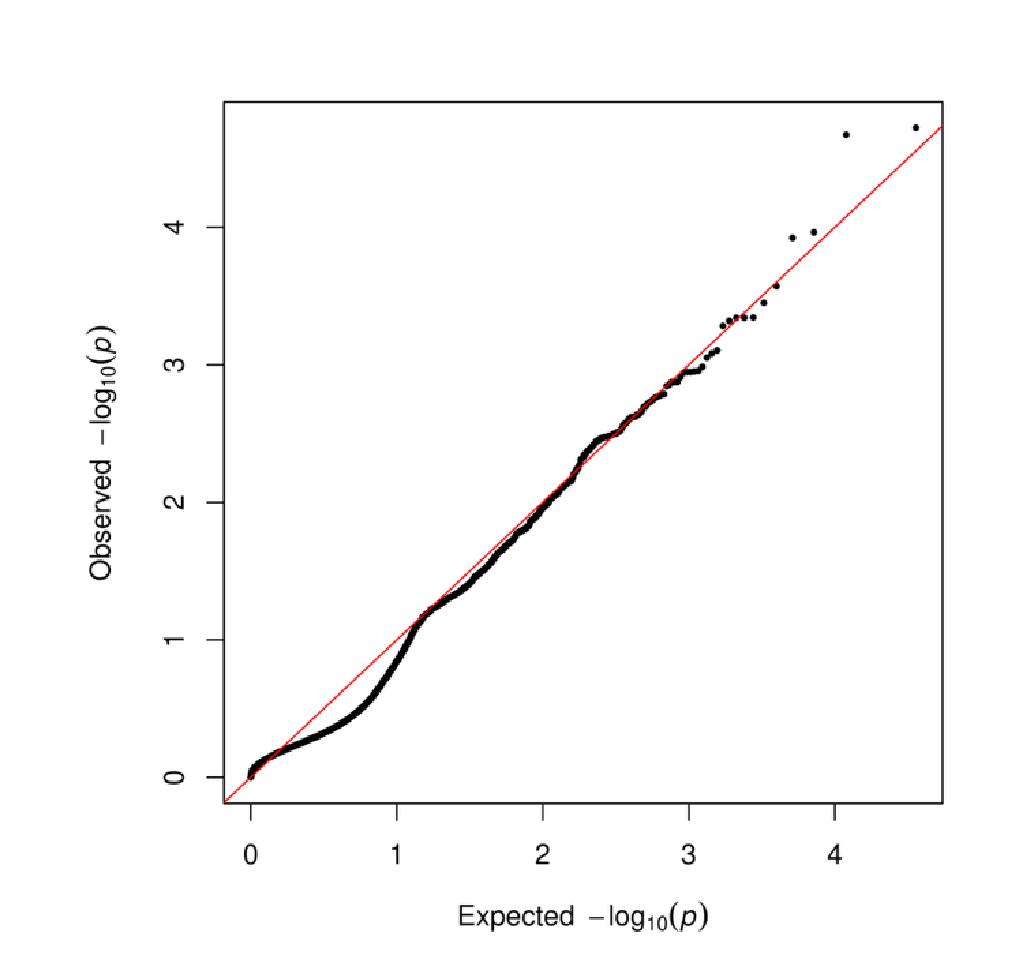

b

c

a

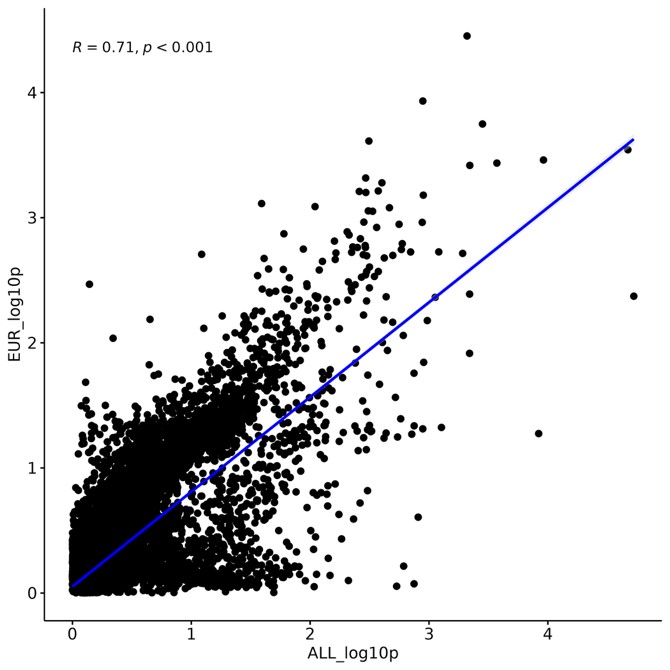

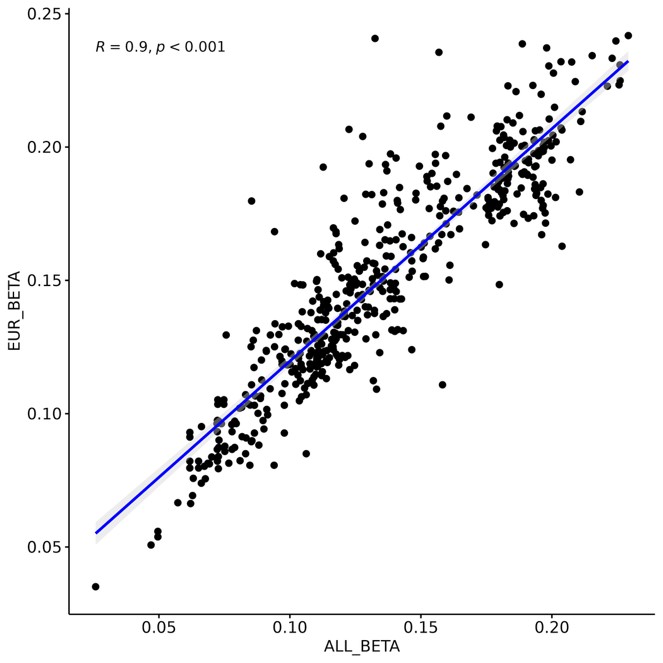

**Supplementary Figure S4.** a) Quantile-quantile (Q-Q) plot of the exome-wide gene-based rare (MAF < 0.01%), likely deleterious missense and loss of function variant analysis for 813 CAKUT cases and 25,205 ancestry-matched controls. Each dot represents a gene. The red line signifies the observed versus the expected -log_10_(*P*) for each gene tested. b) Scatter plot showing the correlation between mixed ancestry and European-only -log_10_(*P*) values. c) Scatter plot showing the correlation between mixed ancestry and European-only BETA effect estimates for variants with *P* < 0.05. *R* is the Spearman correlation coefficient (*rho*).

### Supplementary Figure S5

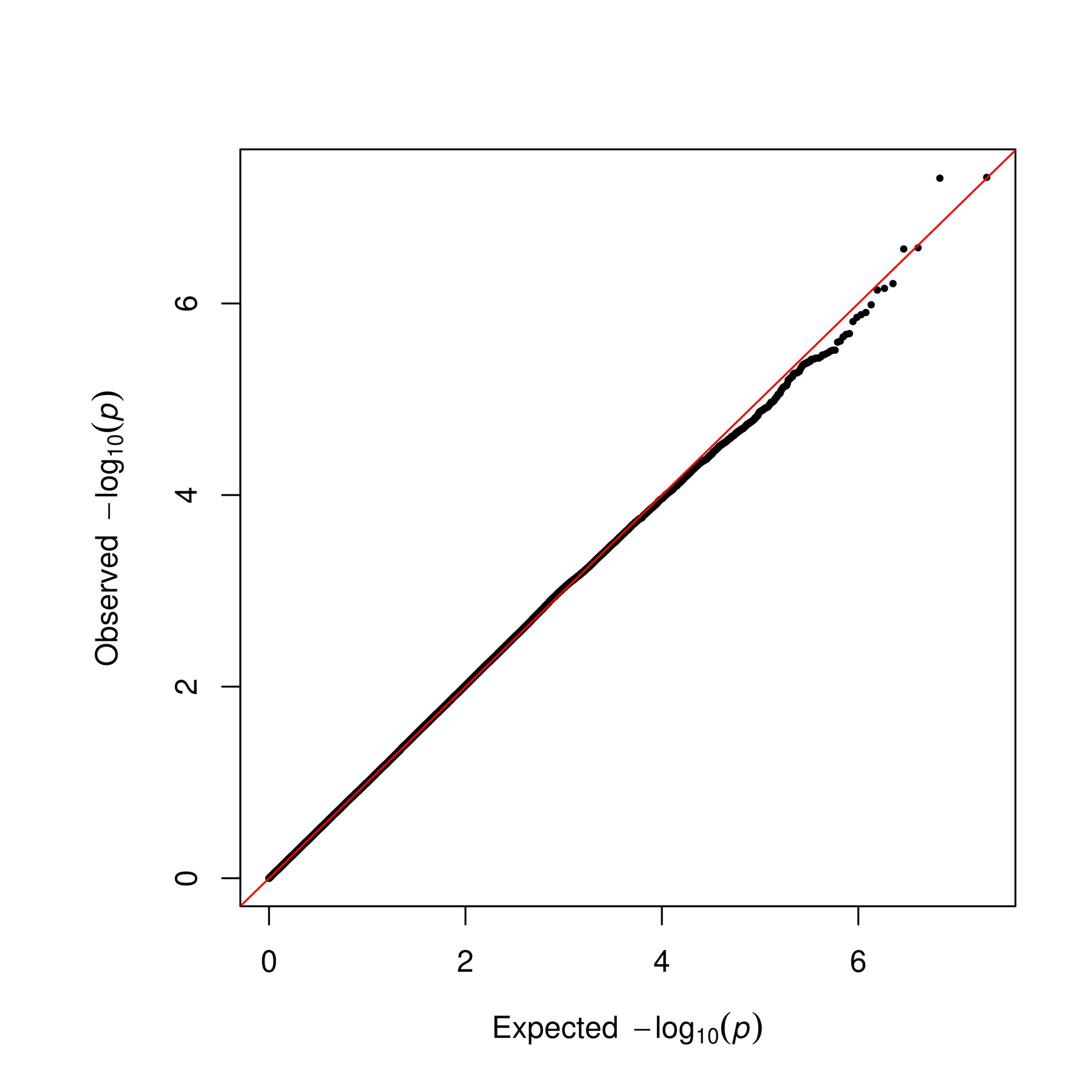

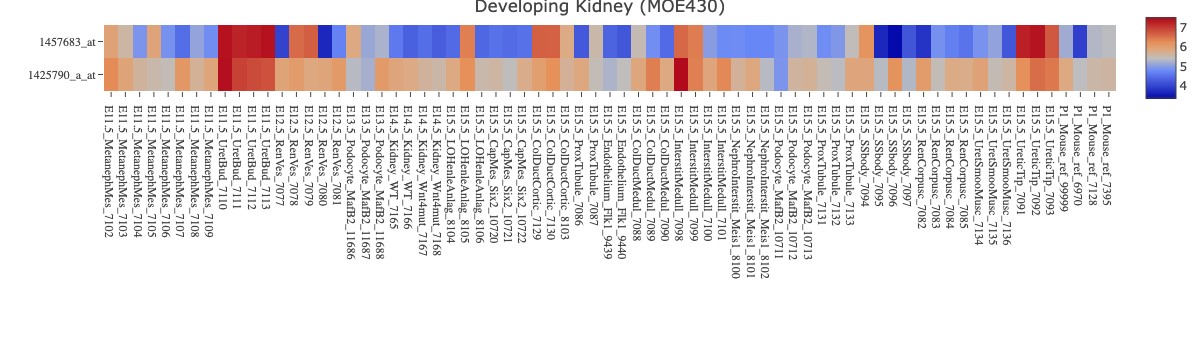

a

b

d

c

λ

1.00

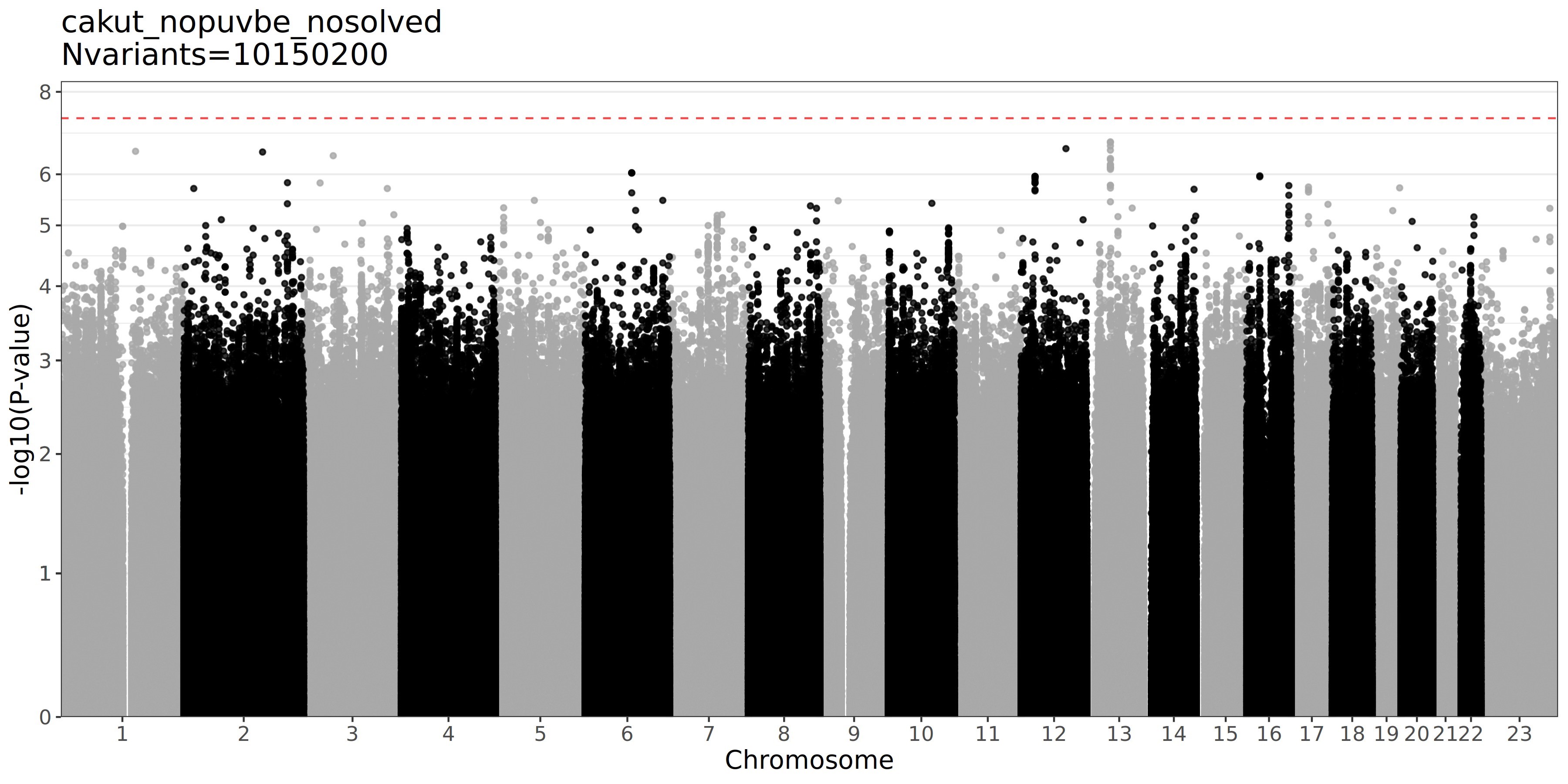

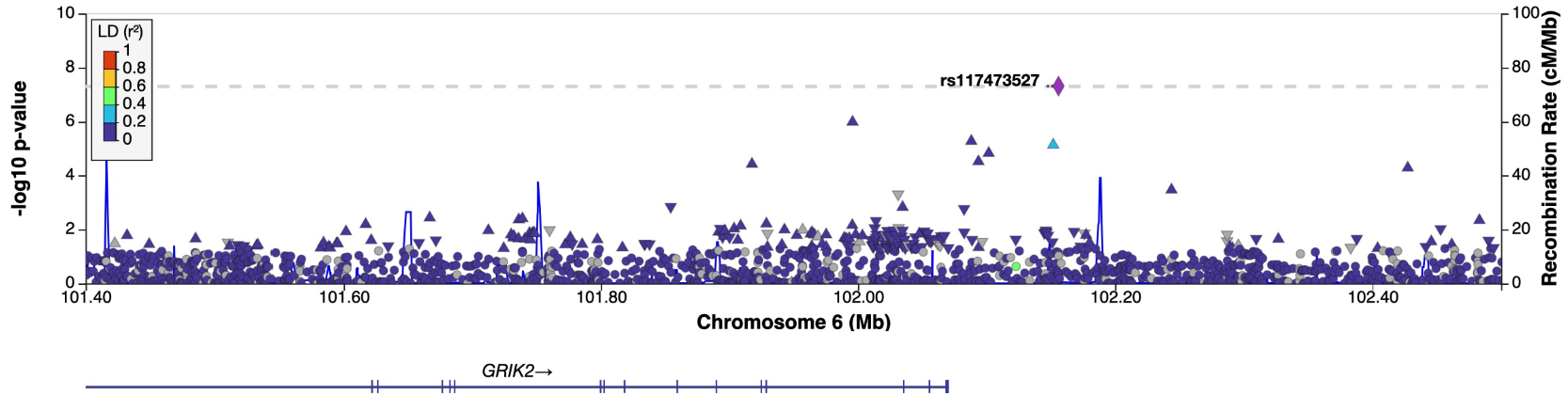

**Supplementary Figure S5. seqGWAS of 813 CAKUT cases and 25,205 ancestry-matched controls**. a) Q-Q

plot displaying the observed versus the expected –log_10_(*P*) for each variant tested. The grey shaded area represents the 95% confidence interval of the null distribution. b) Regional association plot of 6q16.3. Each dot is a variant. Variants are coloured according to their linkage disequilibrium (r^2^) with the lead variant. The red line indicates the genome-wide significance threshold of 5x10^-8^. The gene *GRIK2* is shown against its chromosomal position (GRCh38). c) Heatmap of *Grik2* expression in the murine developing kidney.^S51^ High expression (dark red) is seen in the ureteric bud at E11.5 (rectangle). d)

Manhattan plot of seqGWAS in 551 unsolved CAKUT cases, excluding PUV and bladder exstrophy, and 25,205 controls across 10.2 million variants with MAF > 0.5%. Chromosomal position (GRCh38) is denoted along the x axis and strength of association using a –log_10_(*P*) scale on the y axis. Each dot represents a variant. The red dashed line indicates genome-wide significance at a threshold of *P*=5x10^-8^.

### Supplementary Figure S6

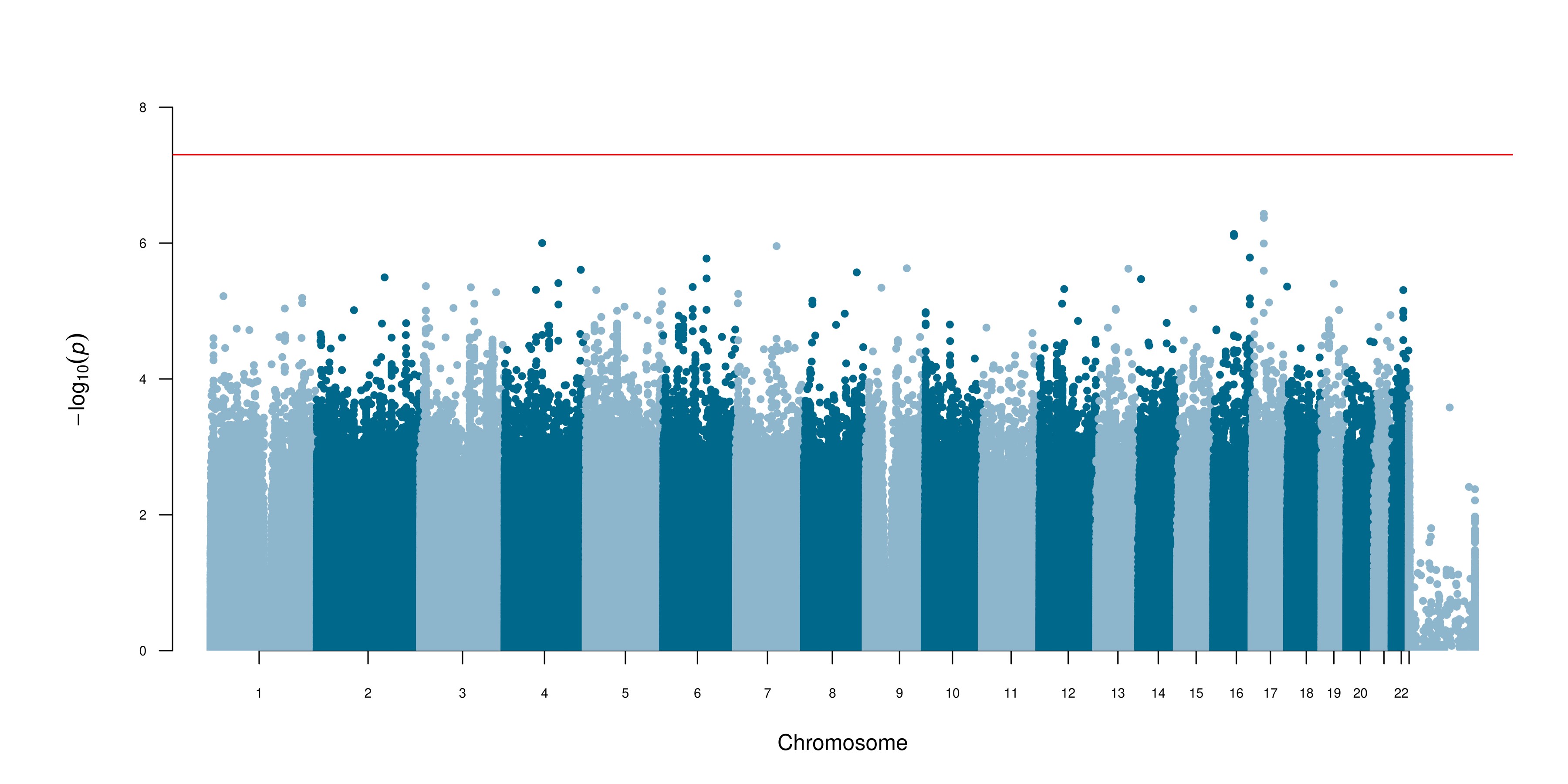

a

b

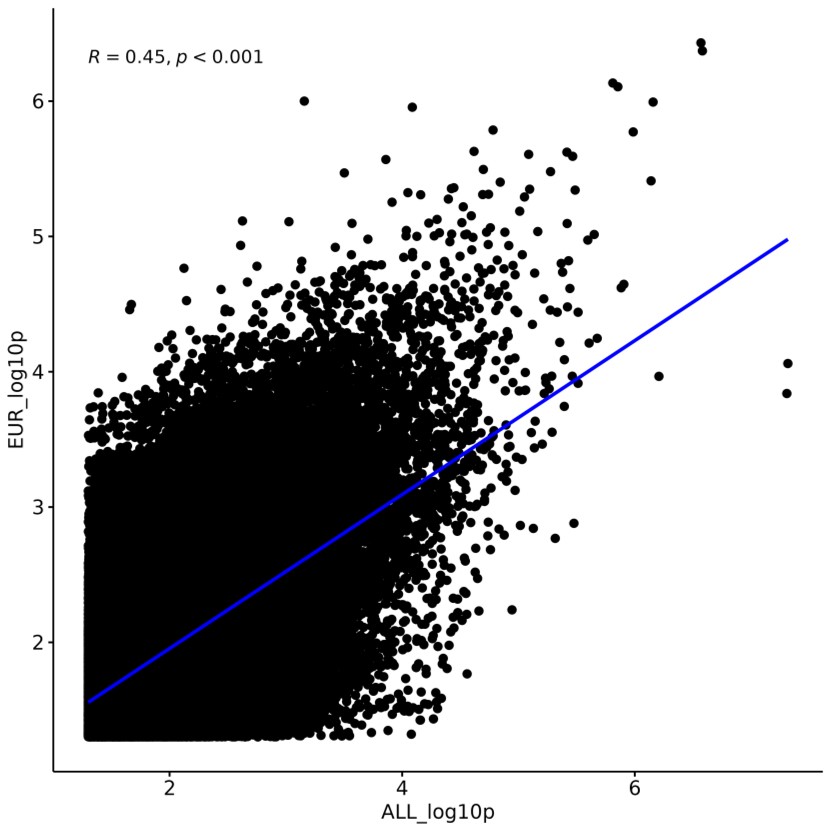

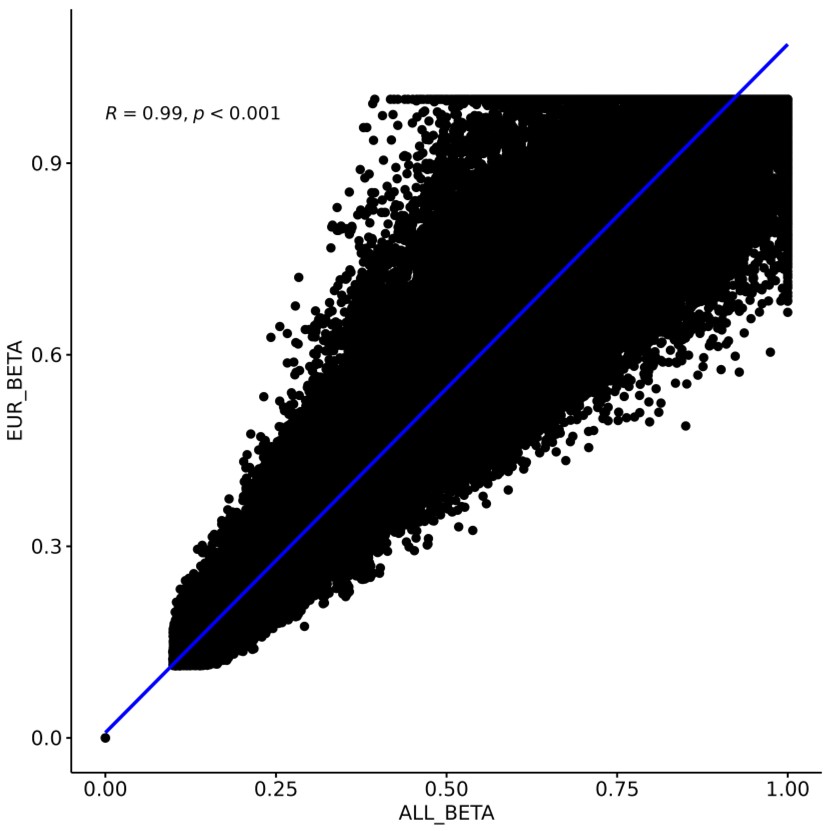

c

d

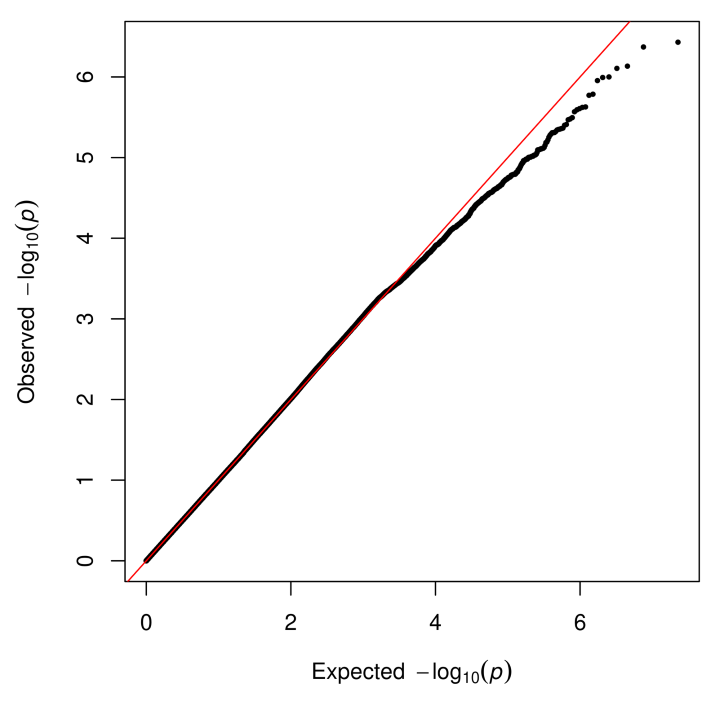

λ

1.00

**Supplementary Figure S6. European-only CAKUT seqGWAS**. a) Manhattan plot of seqGWAS in 623

European CAKUT cases and 20,060 European controls across 11.3 million variants with MAF > 0.5%. Chromosomal position (GRCh38) is denoted along the x axis and strength of association using a –log_10_(*P*) scale on the y axis. Each dot represents a variant. The red dashed line indicates genome-wide significance at a threshold of *P*=5x10^-8^. b) Q-Q plot displaying the observed versus the expected – log_10_(*P*) for each variant tested. The grey shaded area represents the 95% confidence interval of the null distribution. c) Scatter plot showing the correlation between mixed ancestry and European-only -log_10_(*P*) values. d) Scatter plot showing the correlation between mixed ancestry and European-only BETA effect estimates for variants with *P* < 0.05. *R* is the Spearman correlation coefficient (*rho*).

**Supplementary Figure S7**

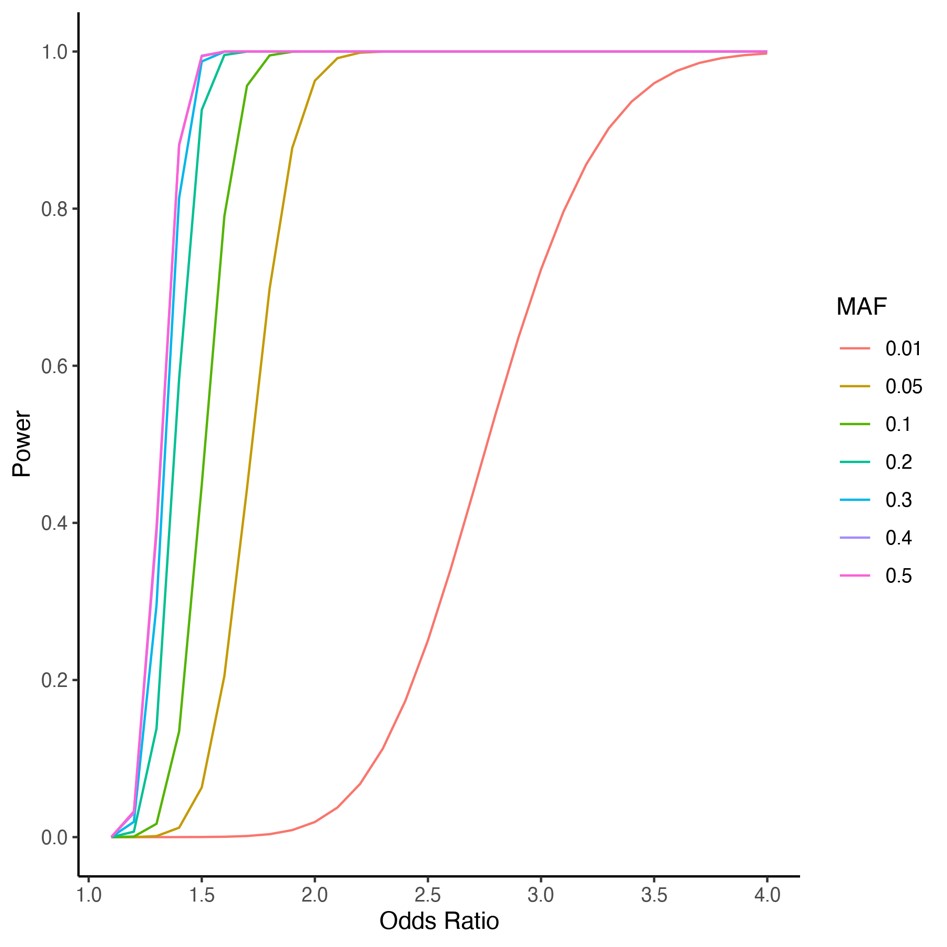

**Supplementary Figure S7. Statistical power for CAKUT GWAS.** Power to detect single variant association under an additive model for 813 CAKUT cases and 25,205 controls at a genome-wide significance threshold of 5x10^-8^. MAF, minor allele frequency.

**Supplementary Figure S8**

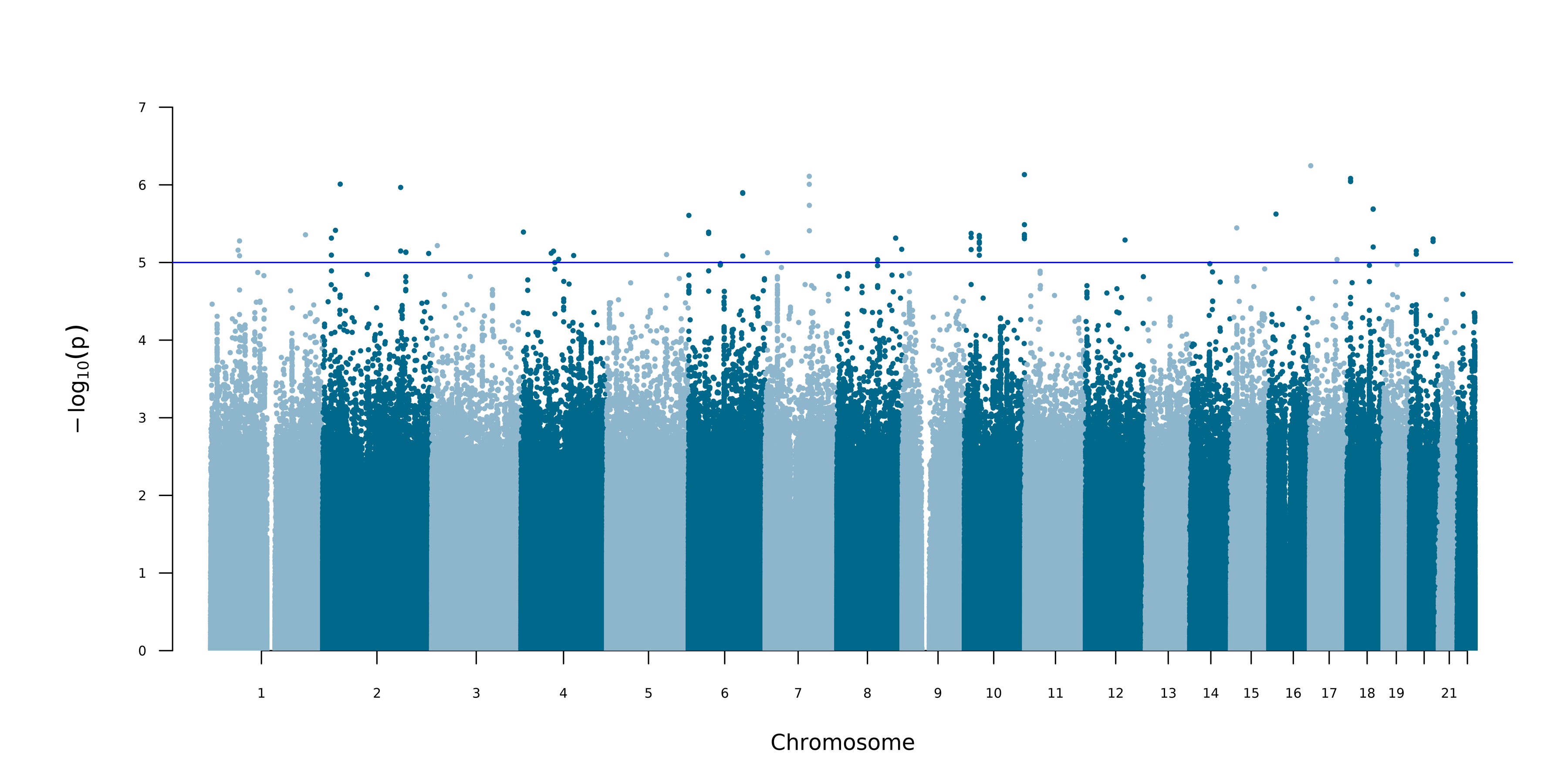

a

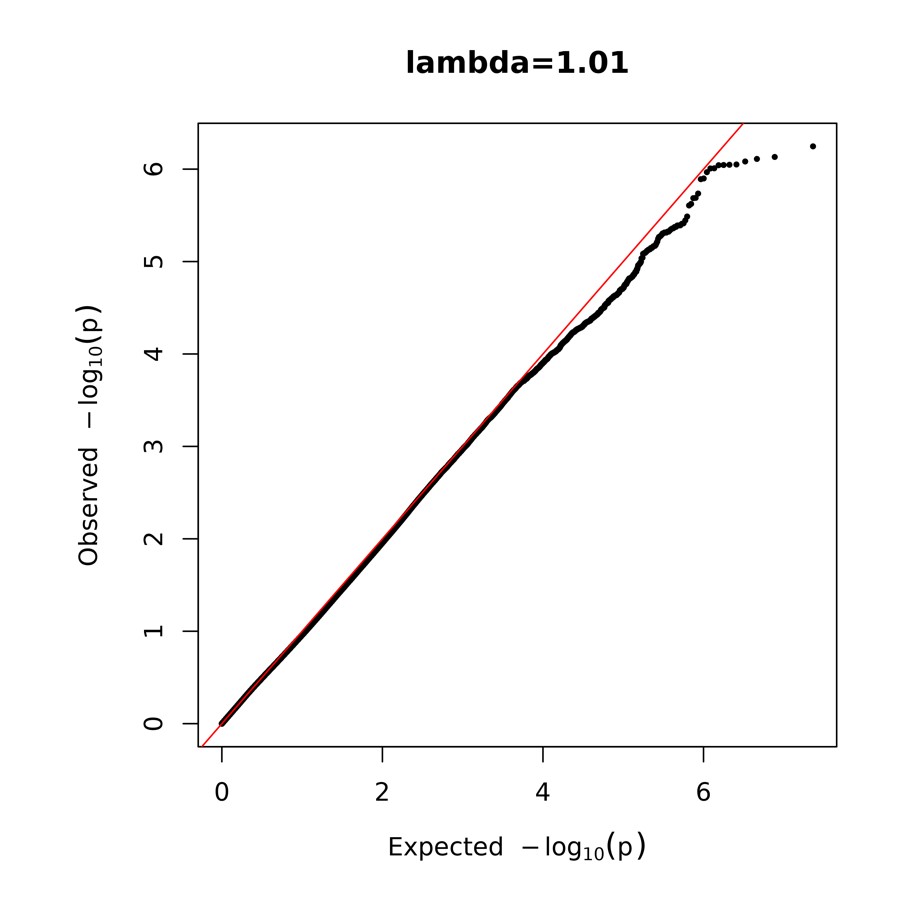

b

**Supplementary Figure S8. Meta-analysis of 727 congenital obstructive uropathy cases and 434,900 controls.** a) Manhattan plot of 11,564,803 variants with MAC > 5. Chromosomal position (GRCh38) is denoted along the x axis and strength of association using a –log_10_(*P*) scale on the y axis. Each dot represents a variant. The blue line indicates suggestive association at a threshold of *P* < 10^-5^. b) Q-Q plot displaying the observed versus the expected –log_10_(*P*) for each variant tested.

**Supplementary Figure S9**

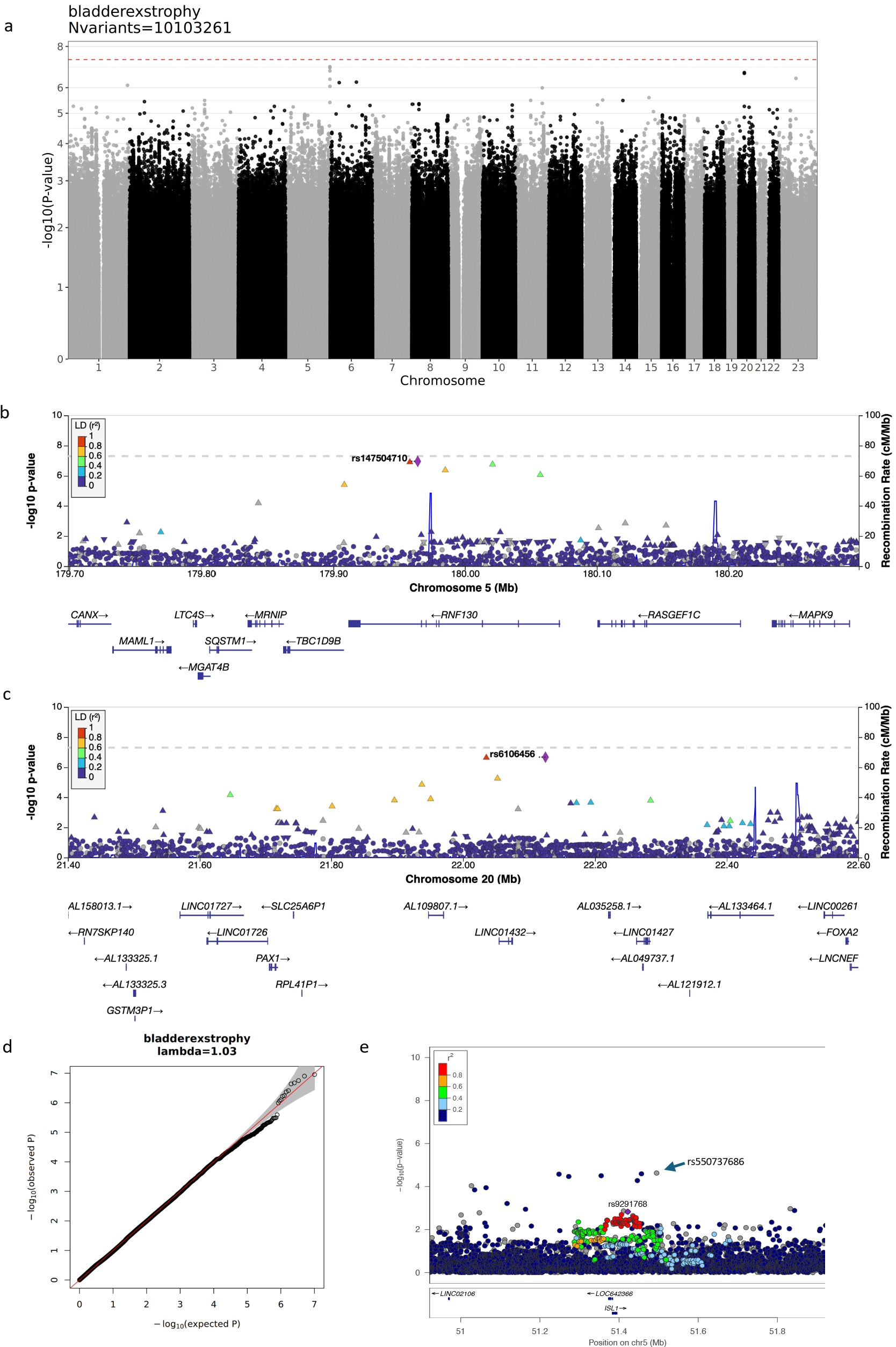

**Supplementary Figure S9. seqGWAS of 97 bladder exstrophy cases and 25,921 ancestry-matched**

**controls.** a) Manhattan plot of 10,103,261 variants with MAF ≥ 0.5%. Chromosomal position (GRCh38) is denoted along the x axis and strength of association using a –log_10_(*P*) scale on the y axis. Each dot represents a variant. The red line indicates the conventional threshold for genome-wide significance (*P* < 5x10^-8^). b) and c) Regional association plots of 5q35.3 and 20p11.22. Each circle is a variant colored according to their linkage disequilibrium (r^2^) with the lead variant. Genes are shown against their chromosomal position (GRCh38). d) Q-Q plot displaying the observed versus the expected –log_10_(*P*) for each variant tested. The grey shaded area represents the 95% confidence interval of the null distribution. e) Regional association plot of previously associated locus at 5q11.1 at higher resolution (variants with MAC > 3). The lead variant at this locus is an indel denoted with a blue arrow. rs9291768 (purple circle) was the lead variant identified in Draaken et al. 2015.^S52^

**Supplementary Figure S10**

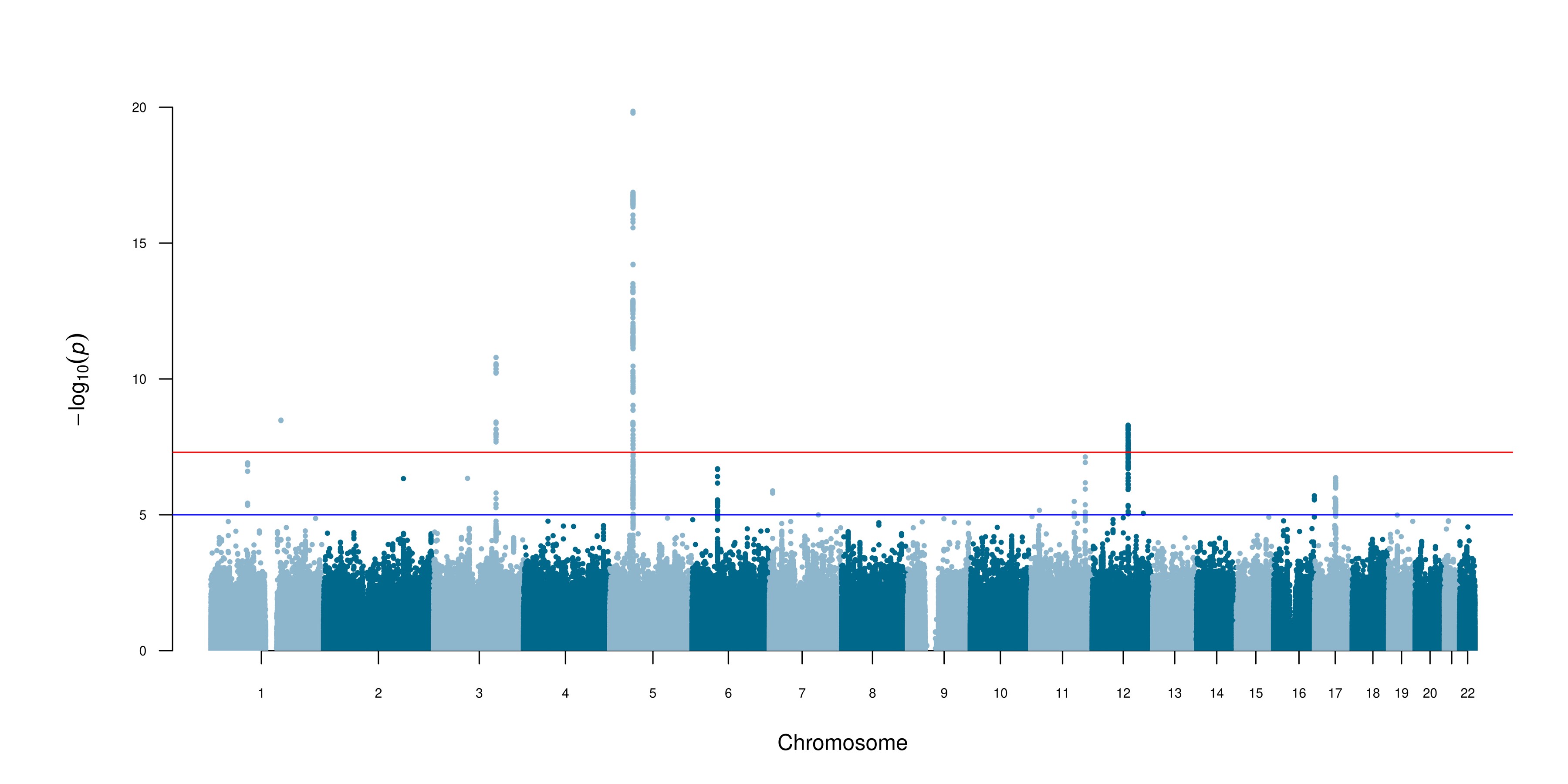

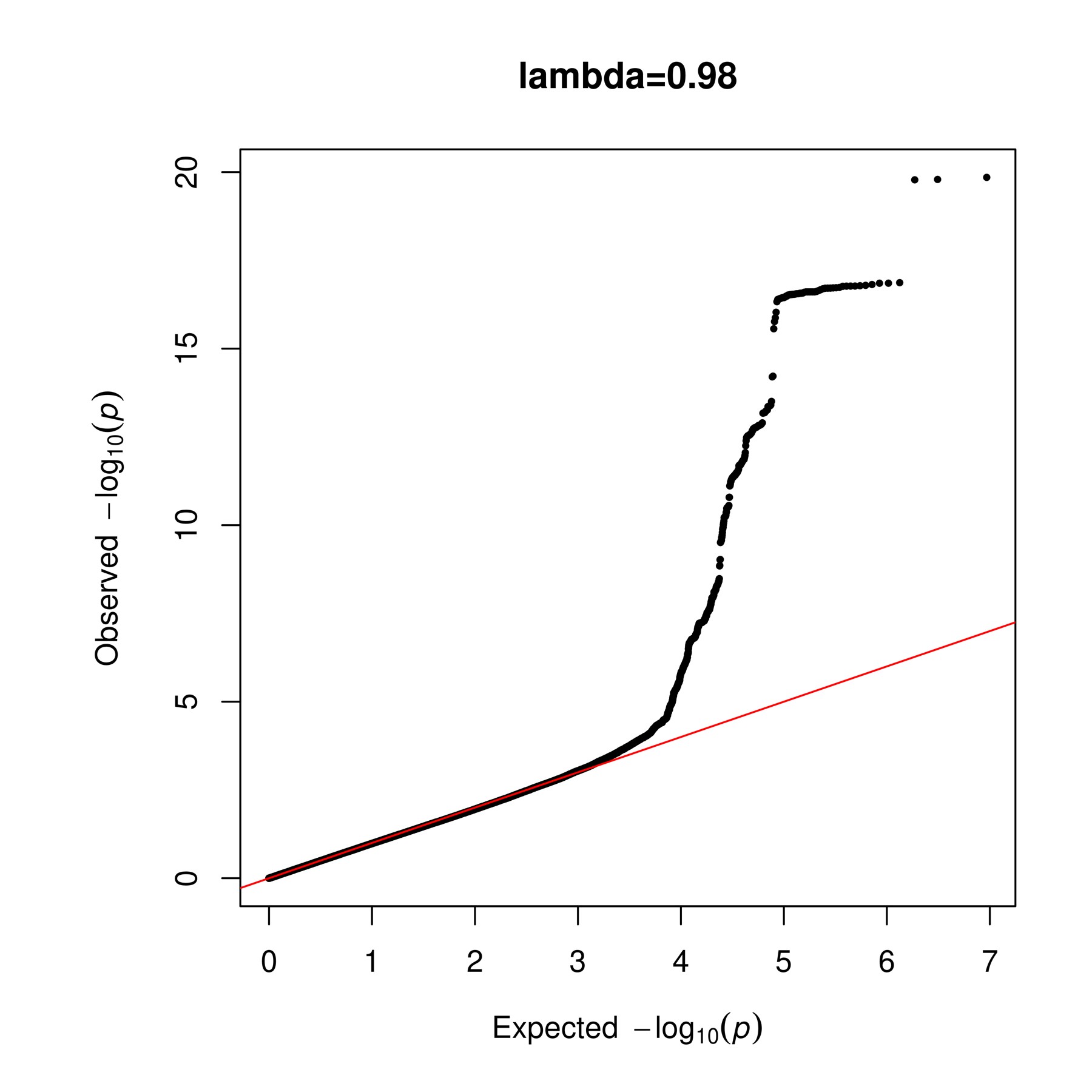

a

b

*TRIM29*

*ISL1*

*ADGRL*

*2*

*SOX14*

*DST*

*EFNA*

*SYT1*

*GOSR2*

*TUBB3*

**Supplementary Figure S10. Meta-analysis of 725 cases with bladder exstrophy and 29,389 controls.** a) Manhattan plot of 4,663,819 variants with MAF ≥ 1%. Chromosomal position (GRCh38) is denoted along the x axis and strength of association using a –log_10_(*P*) scale on the y axis. Each dot represents a variant. The red line indicates the conventional threshold for genome-wide significance (*P* < 5x10^-8^). The blue line indicates suggestive association at a threshold of *P* < 10^-5^. The closest gene to each locus is labelled. b) Q-Q plot displaying the observed versus the expected –log_10_(*P*) for each variant tested.

**Supplementary Figure S11**

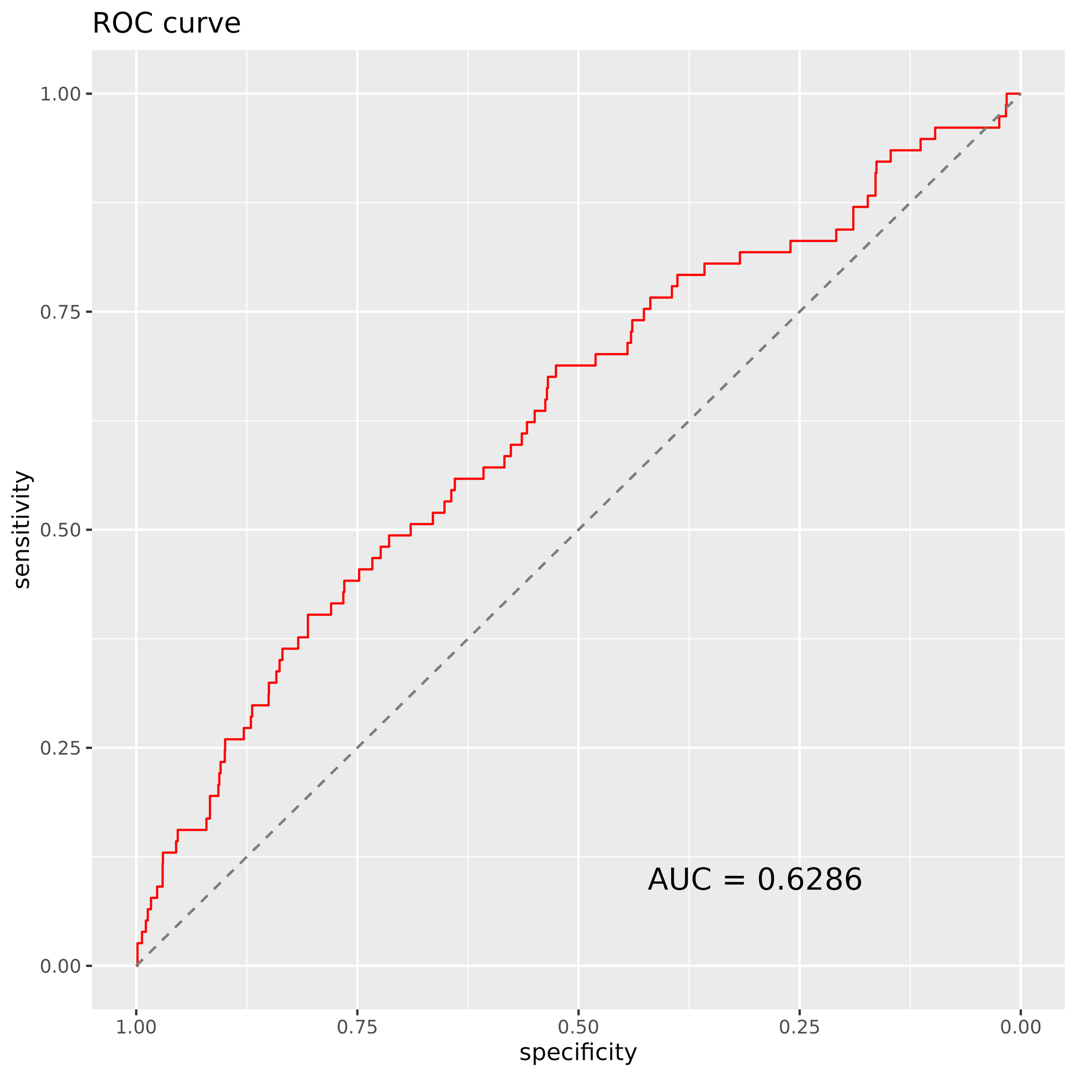

**Supplementary Figure S11. Receiver Operating Characteristic (ROC) curve for PUV-GRS.** Bootstrapped Area Under the Curve (AUC) was 0.63 ± 0.07 (95% confidence interval) when used as a predictor of PUV in a binomial GLM model.

S4. McLaren W, Gil L, Hunt SE, et al. The Ensembl Variant Effect Predictor. *Genome Biol*.

2016;17(1):122.

S5. Rentzsch P, Witten D, Cooper GM, Shendure J, Kircher M. CADD: predicting the deleteriousness of variants throughout the human genome. *Nucleic Acids Res*. 2019;47(D1):D886-D894.

S6. Karczewski KJ, Francioli LC, Tiao G, et al. The mutational constraint spectrum quantified from variation in 141,456 humans. *Nature*. 2020;581(7809):434-443.

S7. Taliun D, Harris DN, Kessler MD, et al. Sequencing of 53,831 diverse genomes from the NHLBI TOPMed Program. *Cold Spring Harbor Laboratory*. Published online March 6, 2019:563866.

doi:10.1101/563866

S8. Danecek P, Bonfield JK, Liddle J, et al. Twelve years of SAMtools and BCFtools. *Gigascience*. 2021;10(2). doi:10.1093/gigascience/giab008

S9. Sadeghi-Alavijeh O, Chan MM, Doctor GT, et al. Quantifying variant contributions in cystic kidney disease using national-scale whole genome sequencing. *J Clin Invest*. Published online August 27, 2024. doi:10.1172/JCI181467

S10. Purcell S, Neale B, Todd-Brown K, et al. PLINK: a tool set for whole-genome association and population-based linkage analyses. *Am J Hum Genet*. 2007;81(3):559-575.

S11. Chang CC, Chow CC, Tellier LC, Vattikuti S, Purcell SM, Lee JJ. Second-generation PLINK: rising to the challenge of larger and richer datasets. *GigaScience*. 2015;4(1). doi:10.1186/s13742-015-0047-8.

S12. Manichaikul A, Mychaleckyj JC, Rich SS, Daly K, Sale M, Chen WM. Robust relationship inference in genome-wide association studies. *Bioinformatics*. 2010;26(22):2867-2873.

S13. Smedley D, Jacobsen JOB, Jäger M, et al. Next-generation diagnostics and disease-gene discovery with the Exomiser. *Nat Protoc*. 2015;10(12):2004-2015.

S14. Verbitsky M, Westland R, Perez A, et al. The copy number variation landscape of congenital anomalies of the kidney and urinary tract. *Nat Genet*. 2019;51(1):117-127.

S15. Ioannidis NM, Rothstein JH, Pejaver V, et al. REVEL: An ensemble method for predicting the pathogenicity of rare missense variants. *Am J Hum Genet*. 2016;99(4):877-885.

S16. Zhou W, Bi W, Zhao Z, et al. SAIGE-GENE+ improves the efficiency and accuracy of set-based rare variant association tests. *Nat Genet*. Published online September 22, 2022. doi:10.1038/s41588022-01178-w

S17. Lee S, Emond MJ, Bamshad MJ, et al. Optimal unified approach for rare-variant association testing with application to small-sample case-control whole-exome sequencing studies. *Am J Hum Genet*.

2012;91(2):224-237.

S18. Zhou W, Nielsen JB, Fritsche LG, et al. Efficiently controlling for case-control imbalance and sample relatedness in large-scale genetic association studies. *Nat Genet*. 2018;50(9):1335-1341.

S19. Dey R, Schmidt EM, Abecasis GR, Lee S. A Fast and Accurate Algorithm to Test for Binary Phenotypes and Its Application to PheWAS. *Am J Hum Genet*. 2017;101(1):37-49.

S20. Turner SD. qqman: an R package for visualizing GWAS results using Q-Q and manhattan plots. *Cold Spring Harbor Laboratory*. Published online May 14, 2014:005165. doi:10.1101/005165

S21. Gogarten SM, Bhangale T, Conomos MP, et al. GWASTools: an R/Bioconductor package for quality control and analysis of genome-wide association studies. *Bioinformatics*. 2012;28(24):3329-3331.

S22. Pruim RJ, Welch RP, Sanna S, et al. LocusZoom: regional visualization of genome-wide association scan results. *Bioinformatics*. 2010;26(18):2336-2337. doi:10.1093/bioinformatics/btq419

S23. de Leeuw CA, Mooij JM, Heskes T, Posthuma D. MAGMA: generalized gene-set analysis of GWAS data. *PLoS Comput Biol*. 2015;11(4):e1004219.

S24. Liberzon A, Birger C, Thorvaldsdóttir H, Ghandi M, Mesirov JP, Tamayo P. The Molecular Signatures Database (MSigDB) hallmark gene set collection. *Cell Syst*. 2015;1(6):417-425.

S25. Nishimura D. BioCarta. *Biotech Software & Internet Report*. 2001;2(3):117-120.

S26. Du J, Yuan Z, Ma Z, Song J, Xie X, Chen Y. KEGG-PATH: Kyoto encyclopedia of genes and genomesbased pathway analysis using a path analysis model. *Mol Biosyst*. 2014;10(9):2441-2447.

S27. Jassal B, Matthews L, Viteri G, et al. The reactome pathway knowledgebase. *Nucleic Acids Res*.

2020;48(D1):D498-D503.

S28. Martens M, Ammar A, Riutta A, et al. WikiPathways: connecting communities. *Nucleic Acids Res*.

2021;49(D1):D613-D621.

S29. Schaefer CF, Anthony K, Krupa S, et al. PID: the Pathway Interaction Database. *Nucleic Acids Res*.

2009;37(Database issue):D674-9.

S30. The Gene Ontology resource: enriching a GOld mine. *Nucleic Acids Res*. 2021;49(D1):D325-D334.

S31. Kurki MI, Karjalainen J, Palta P, et al. FinnGen provides genetic insights from a well-phenotyped isolated population. *Nature*. 2023;613(7944):508-518.

S32. Willer CJ, Li Y, Abecasis GR. METAL: fast and efficient meta-analysis of genomewide association scans. *Bioinformatics*. 2010;26(17):2190-2191.

S33. Mingardo E, Beaman G, Grote P, et al. A genome-wide association study with tissue transcriptomics identifies genetic drivers for classic bladder exstrophy. *Commun Biol*. 2022;5(1):1203.

S34. Kichaev G, Yang WY, Lindstrom S, et al. Integrating functional data to prioritize causal variants in statistical fine-mapping studies. *PLoS Genet*. 2014;10(10):e1004722.

S35. 1000 Genomes Project Consortium, Auton A, Brooks LD, et al. A global reference for human genetic variation. *Nature*. 2015;526(7571):68-74.

S36. Frankish A, Diekhans M, Ferreira AM, et al. GENCODE reference annotation for the human and mouse genomes. *Nucleic Acids Res*. 2019;47(D1):D766-D773.

S37. Weinstock GM, Wilson RK, Gibbs RA, Kent WJ. Evolutionarily conserved elements in vertebrate, insect, worm, and yeast genomes. *Genome*. Published online 2005. https://genome.cshlp.org/content/15/8/1034.short

S38. ENCODE Project Consortium, Moore JE, Purcaro MJ, et al. Expanded encyclopaedias of DNA elements in the human and mouse genomes. *Nature*. 2020;583(7818):699-710.

S39. Krietenstein N, Abraham S, Venev SV, et al. Ultrastructural Details of Mammalian Chromosome Architecture. *Mol Cell*. 2020;78(3):554-565.e7.

S40. Yang J, Bakshi A, Zhu Z, et al. Genetic variance estimation with imputed variants finds negligible missing heritability for human height and body mass index. *Nat Genet*. 2015;47(10):1114-1120.

S41. Falconer DS. The inheritance of liability to certain diseases, estimated from the incidence among relatives. *Ann Hum Genet*. 1965;29(1):51-76.

S42. Public Health England. *National Congenital Anomaly and Rare Disease Registration Service.*

*Congenital Anomaly Statistics 2019*. Dandy Booksellers; 2022.

S43. Speed D, Balding DJ. SumHer better estimates the SNP heritability of complex traits from summary statistics. *Nat Genet*. 2019;51(2):277-284.

S44. Raney BJ, Barber GP, Benet-Pagès A, et al. The UCSC Genome Browser database: 2024 update.

*Nucleic Acids Res*. 2024;52(D1):D1082-D1088.

S45. Finucane HK, Bulik-Sullivan B, Gusev A, et al. Partitioning heritability by functional annotation using genome-wide association summary statistics. *Nat Genet*. 2015;47(11):1228-1235.

S46. Ge T, Chen CY, Ni Y, Feng YCA, Smoller JW. Polygenic prediction via Bayesian regression and continuous shrinkage priors. *Nat Commun*. 2019;10(1):1776.

S47. Lambert SA, Wingfield B, Gibson JT, et al. Enhancing the Polygenic Score Catalog with tools for score calculation and ancestry normalization. *Nat Genet*. 2024;56(10):1989-1994.

S48. Cavalli-Sforza LL. The Human Genome Diversity Project: past, present and future. *Nat Rev Genet*.

2005;6(4):333-340.

S49. Lee, S. H., Goddard, M. E., Wray, N. R., & Visscher, P. M. (2012). A better coefficient of determination for genetic profile analysis. *Genetic Epidemiology*, *36*(3), 214–224.

S50. van Rooij IALM, van der Zanden LFM, Bongers EMHF, et al. AGORA, a data- and biobank for birth defects and childhood cancer. *Birth Defects Res A Clin Mol Teratol*. 2016;106(8):675-684.

S51. Harding SD, Armit C, Armstrong J, et al. The GUDMAP database--an online resource for genitourinary research. *Development*. 2011;138(13):2845-2853.

S52. Draaken M, Knapp M, Pennimpede T, et al. Genome-wide association study and meta-analysis identify ISL1 as genome-wide significant susceptibility gene for bladder exstrophy. *PLoS Genet*.

2015;11(3):e1005024.
